# Preventing childhood obesity primary schools: a realist review from UK perspective

**DOI:** 10.1101/2021.04.15.21255539

**Authors:** Sharea Ijaz, James Nobles, Laura Johnson, Theresa Moore, Jelena Savović, Russell Jago

## Abstract

**Objective:** to identify contextual and mechanistic factors associated with outcomes of school-based obesity prevention interventions which may be implementable in UK primary schools.

**Design:** realist synthesis following REMESES guidance. We developed an initial programme theory through expert consensus and stakeholder input and refined it with data from included studies to produce a final programme theory including all context-mechanism-outcome configurations.

**Setting:** primary schools

**Participants:** children aged 4-12

**Interventions:** included in the 2019 Cochrane review on prevention of childhood obesity

**Outcomes:** BMIz

**Results:** We included 24 studies comprised of 71 documents in our synthesis. We found that contextual factors such as baseline BMIz affects intervention mechanisms variably, while girls, older children and those with higher parental education consistently benefitted more from school-based interventions. Key mechanisms associated with beneficial effect were sufficient intervention dose, environmental modification, and the intervention components working together as a whole. Education alone did not produce favourable outcomes.

**Conclusions:** Interventions should go beyond education and incorporate a sufficient dose to trigger change in BMIz. Contextual factors deserve consideration when commissioning interventions to avoid widening health inequalities.

## 1. Background

The world has witnessed a rapid increase in the prevalence of childhood obesity in the last three decades. UK data indicate that 20% of children aged 4-5 years and one third of children aged 10-11 years currently have overweight or obesity – as classified via use of standardised body mass index (BMIz).^1^ Strategies to prevent excessive weight gain are therefore needed.

Obesity is now widely accepted as a function of a complex and obesogenic society and system.^2-4^ Population-levels of obesity are known to be the product of many interrelated and interdependent factors,^5^ and in response researchers, practitioners and policy makers have started to call for the implementation of a systems approach. These approaches acknowledge that many different sectors, organisations, communities, families and individuals need to come together to systematically address the root causes of obesity.^2^ Given that children spend approximately 25% of their waking hours in schools, and the important role that schools play within society, they serve as a key setting for obesity prevention efforts.^6 7^ Although, schools cannot be expected to prevent childhood obesity on their own, they make up an important part of the system where interventions can go beyond targeting individual responsibility.

The latest Cochrane review ^8^ found that school-based obesity prevention interventions can achieve small changes in BMIz over a school year. However, as interventions varied widely in design and degree of success, the review does not highlight to public health professionals which intervention features work best, for whom, and in what contexts. Realist reviews can help answer these questions by identifying contexts and mechanisms associated with intervention outcomes. ^9 10 11^

The aim of this realist review was to identify, and understand, the contextual and mechanistic factors associated with the outcome of school-based obesity prevention studies included in the Cochrane review of Brown et al.^8^, which may be implemented within UK primary schools.

## 2. Methods

We carried out a realist review underpinned by the RAMESES guidance and the existing realist reviews in similar fields. ^10 11^ The study was registered with PROSPERO in July 2019 (CRD42019142192).^12^

### 2.1 Development of a programme theory

We developed an initial programme theory (Figure 1) using our team expertise in obesity prevention, and intervention development and evaluation.

**Figure 1.**
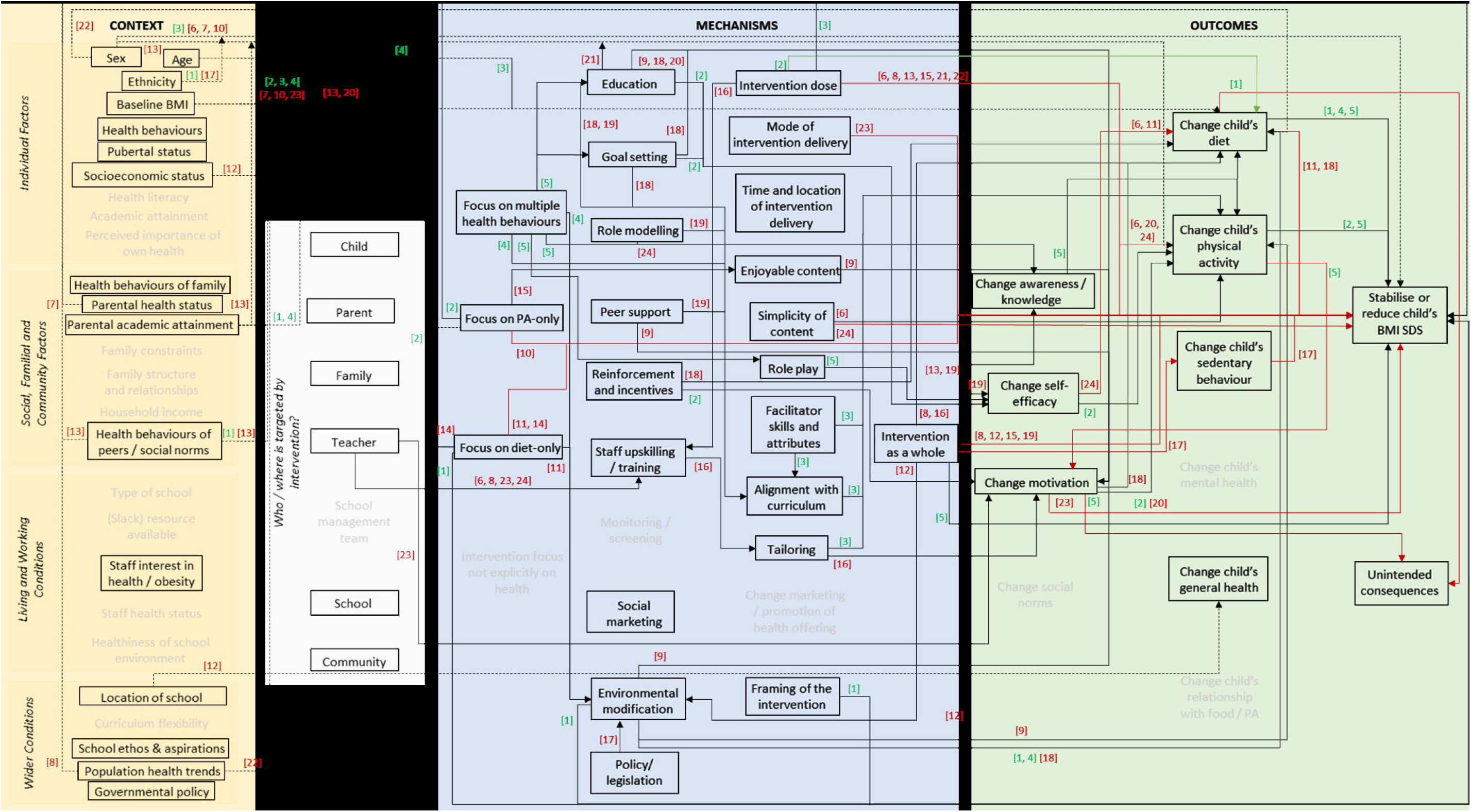
The initial programme theory.

Patient and Public Involvement: We sought external stakeholder consultation^13 14^ – via an online survey - to facilitate our understanding of the UK primary school contexts, and what stakeholders (school staff, management and organisations that work with primary schools) consider important.

The initial theory outlined the contextual and mechanistic factors that may be associated with change in BMIz among children aged 4-12 years old exposed to a primary school-based intervention. This programme theory was further developed and refined with stakeholder input and data from included studies over the course of the review in an iterative manner. Supporting information (section 1) illustrates how the programme theory evolved.

### 2.2 Inclusion / exclusion criteria

Our sample frame was the recent Cochrane review “Interventions for Preventing Obesity in Children” which included 153 studies.^8^ We included studies which met the following criteria: conducted in primary schools; included children aged 4-12 years; interventions aimed to prevent obesity; and presented mean BMIz as an outcome.

### 2.3 Data extraction (selection and coding)

Two reviewers (SI, JN) assessed all studies included in the Cochrane review to determine if a study met our inclusion criteria. Data were extracted into a standardised template (see supporting information section 2) which evolved as review progressed. Whenever we identified a new context or mechanism during data extraction, we added these to data extraction forms and then revisited the previously extracted studies to ensure data were not overlooked. Over repeated rounds, and along with input from topic experts on the team (JN, LJ and RJ), we reached consensus over the coding for all extracted texts.

### 2.4 Rigour assessment

We operationalised rigour assessment into a four-point scale based on the RAMESES definition of rigour^15^ which are presented below. We employed risk of bias^16^ judgements for the outcome as reported in the Cochrane review. ^8^ These decisions were made case by case and agreed between two reviewers (SI, JN) (see example in supporting information section 3).

The four categories of rigour for studies were:

- Highly rigorous data (++): Arguments /data for the CMOs are appropriate (underpinned with theory and data), and study was at low risk of bias for our outcome.
- Rigorous data (+): Arguments/ data presented are appropriate for CMOs, and study is not at low risk of bias for our outcome.
- Unclear rigour of data (?): No or weak arguments/ data presented for CMOs, irrespective of whether study is at low risk of bias for our outcome.
- Data not rigorous (-): Contrary or unreliable arguments/ data presented, irrespective of whether study is at low risk of bias for our outcome.

### 2.5 Data synthesis

Synthesis was a two-stage process. We first presented data on the CMO configurations at study level. Thus, producing a programme theory diagram for each study describing its CMO configurations. Then, for stage 2, we collated the CMO configurations from each study into a single, synthesised programme theory diagram (Figure 2).

**Figure 2.**
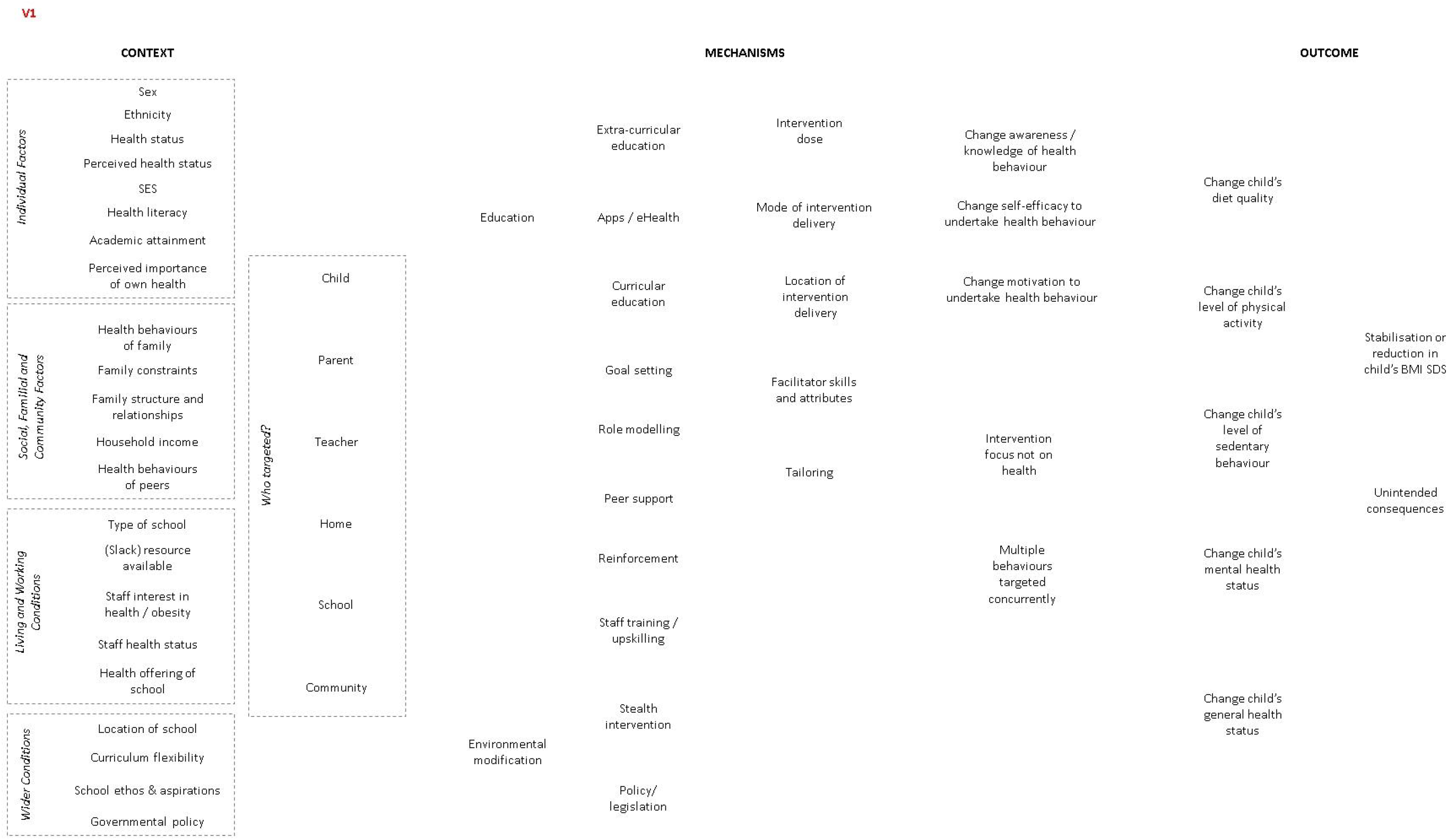
Final programme theory showing CMOs from all included studies. Dotted black lines indicate which contexts affected which outcomes. Continuous black lines from mechanism to outcomes indicate a favourable change (e.g. improved PA levels) while continuous red lines indicate of a favourable change (e.g. no difference in PA levels or an unfavourable change (e.g. lower PA levels). [] bracketed numbers underneath the lines indicate respective studies for that CMO line. Green brackets refer to studies that found a favourable BMIz change (effective studies) and red refer to those that did not ffective studies): 1=deRuyter 2012;2=Khan 2014; 3= Li 2010; 4=Marcus 2009; 5=Spiegel 2006; 6=Fairclough 2013; 7=Cao 2015; 8=Sahota 2001; 9=Gutin 2008; 10=Lazaar 2007; 11=Damsgaard 2014; 12=Rush 2; 13=Grydeland 2014; 14= James 2004; 15=Meng 2013; 16=Rosario 2012; 17=Foster 2008; 18= Muckelbauer 2010; 19=Santos 2014; 20=Siegrist 2013; 21=Williamson 2012; 22=Herscovici 2013; 23=Johnston 3; 24= Kipping 2014.

We also summarised data reported on costs and sustainability of the interventions (supporting information section 4), as stakeholders considered these important.

### 2.6 Analysis of subgroups or subsets

We present programme theories for effective (defined as statistically significant BMIz change favouring intervention as seen in the Cochrane review) and ineffective interventions in supporting information section 5. We also synthesised studies with rigorous data alone to see any differences from main synthesis (see supporting information section 5).

## 3. Results

All 153 studies included in the Cochrane review were assessed at abstract stage against our inclusion criteria. Of these, 29 studies met the criteria and were assessed in full texts (81 documents). Five studies (10 documents) were excluded at this stage as these were set entirely outside of the school ^17-19^ or did not involve primary school aged children.^20 21^ Thus 24 studies^22-45^ (71 documents) were included in this realist review. See supporting information section 6 for study flow and lists of excluded and included study documents.

### 3.2 Included study characteristics

See details of studies and extracted data in table 1.

**Table 1.**
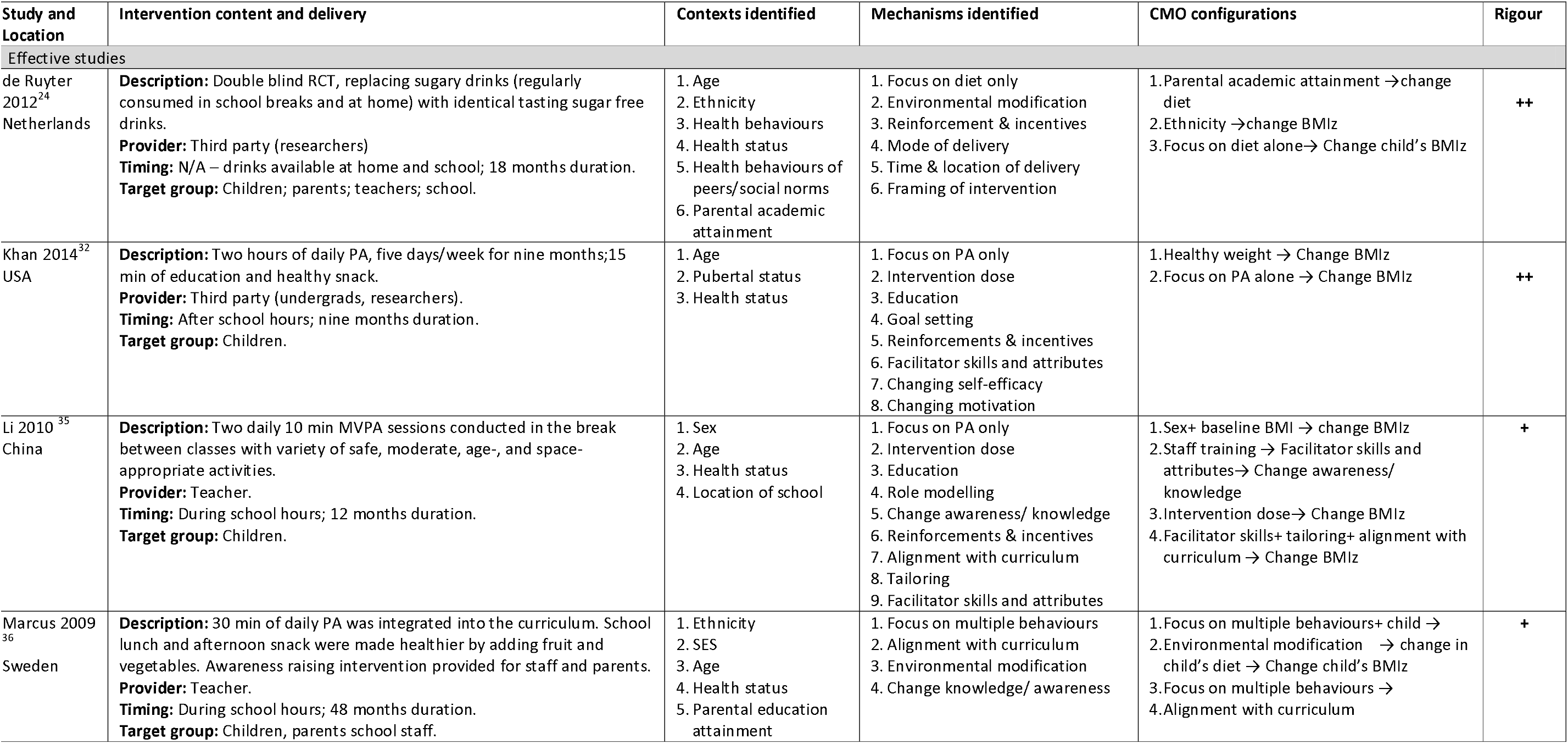
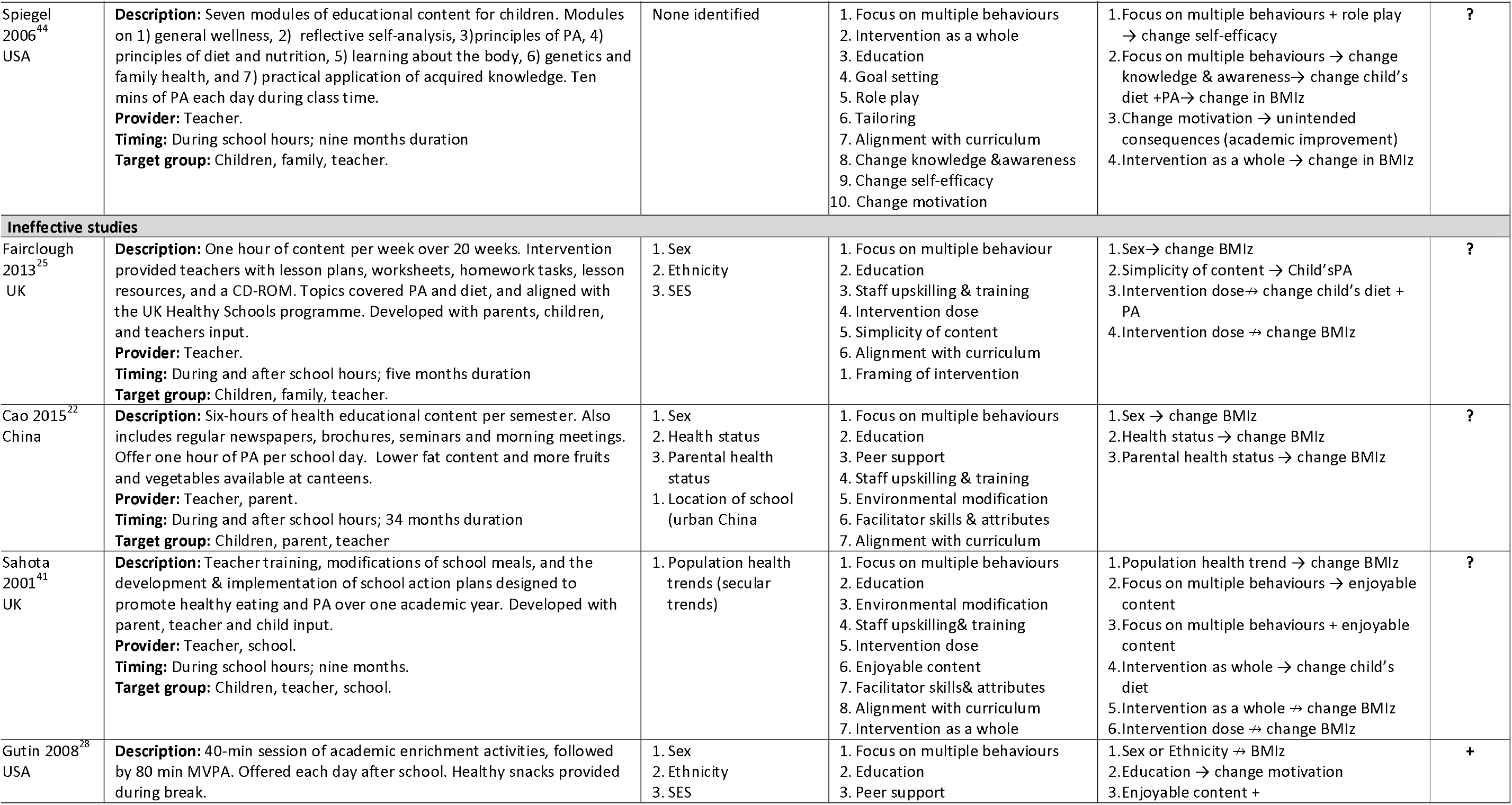
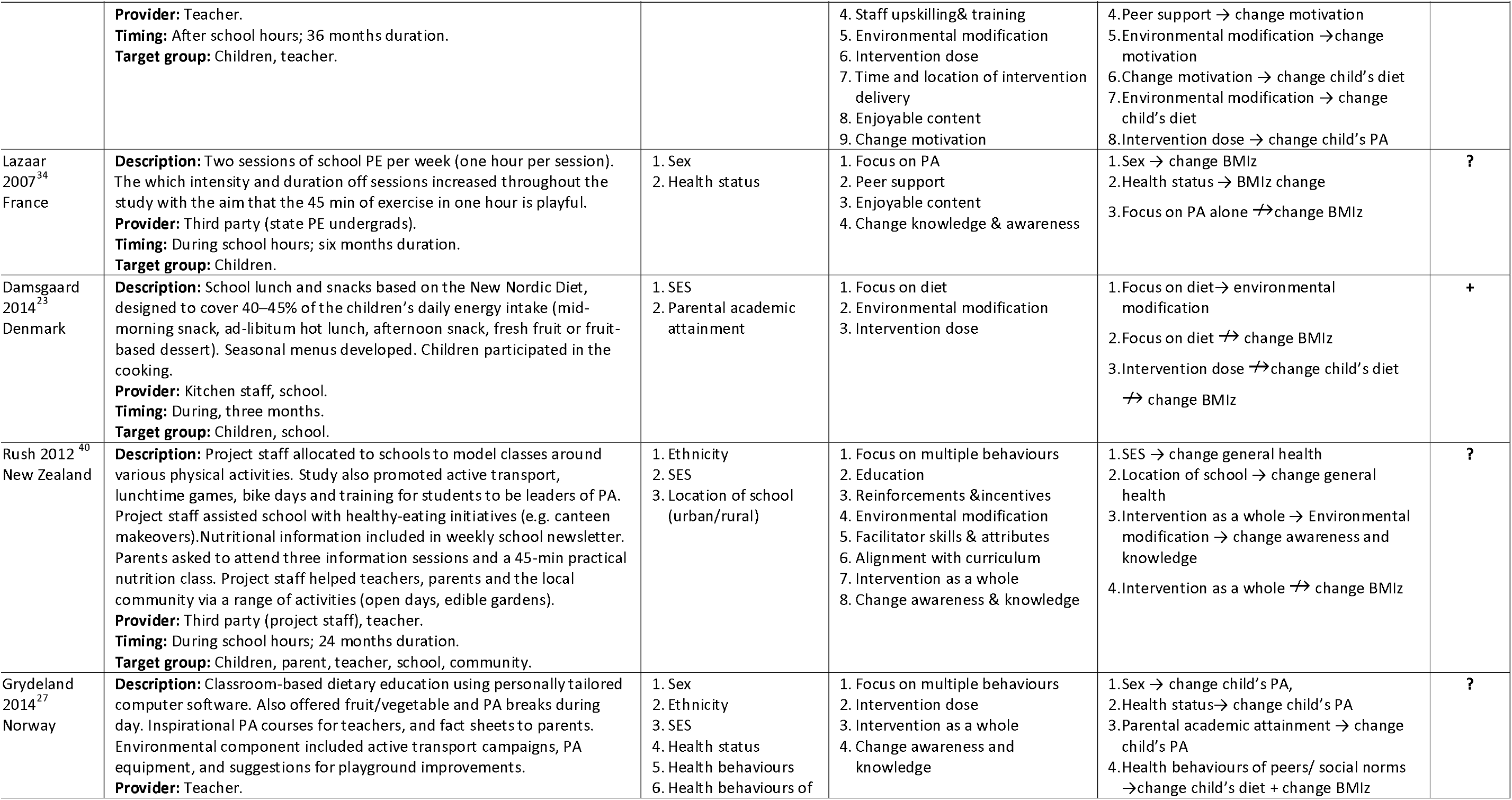
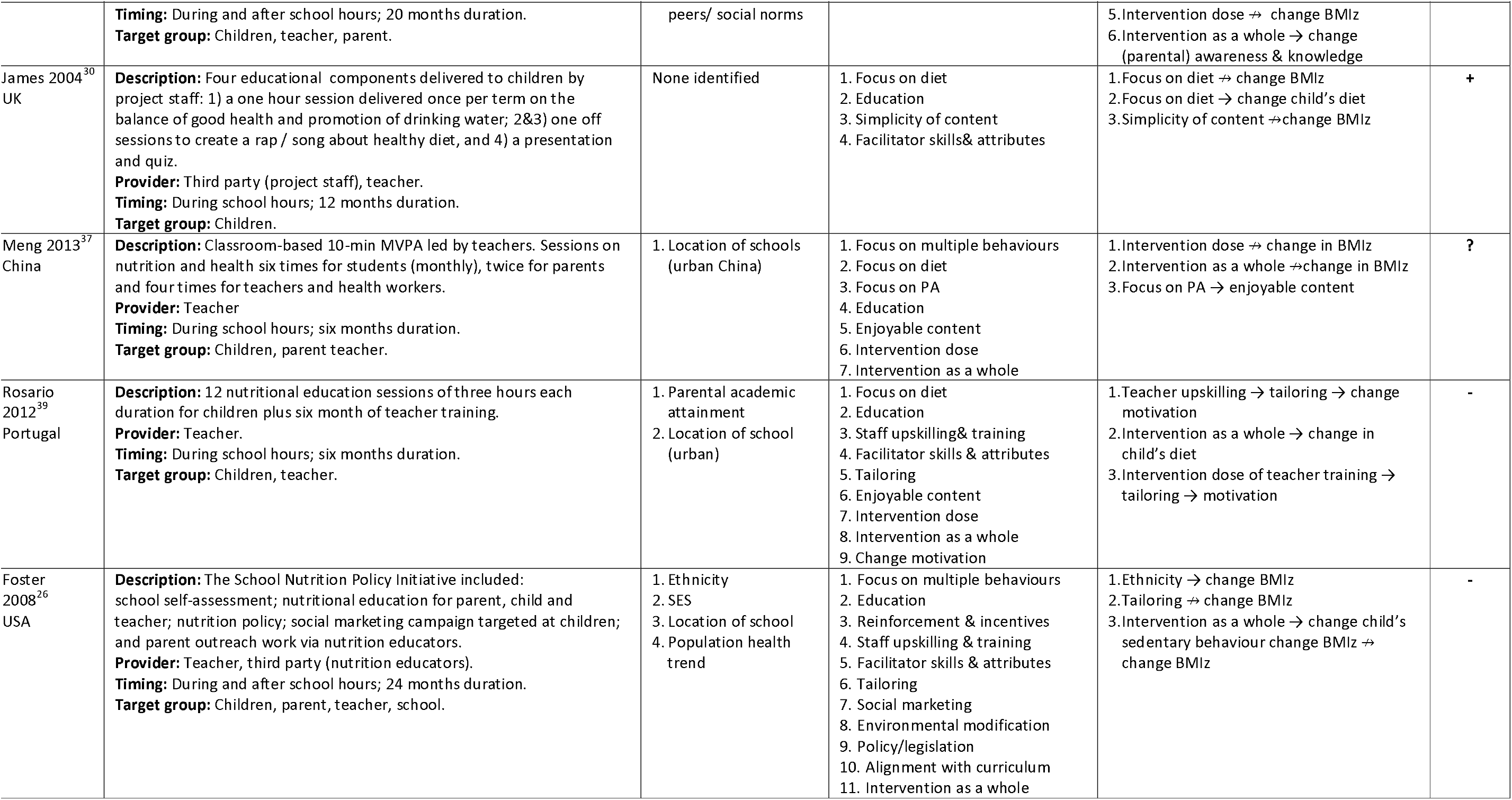
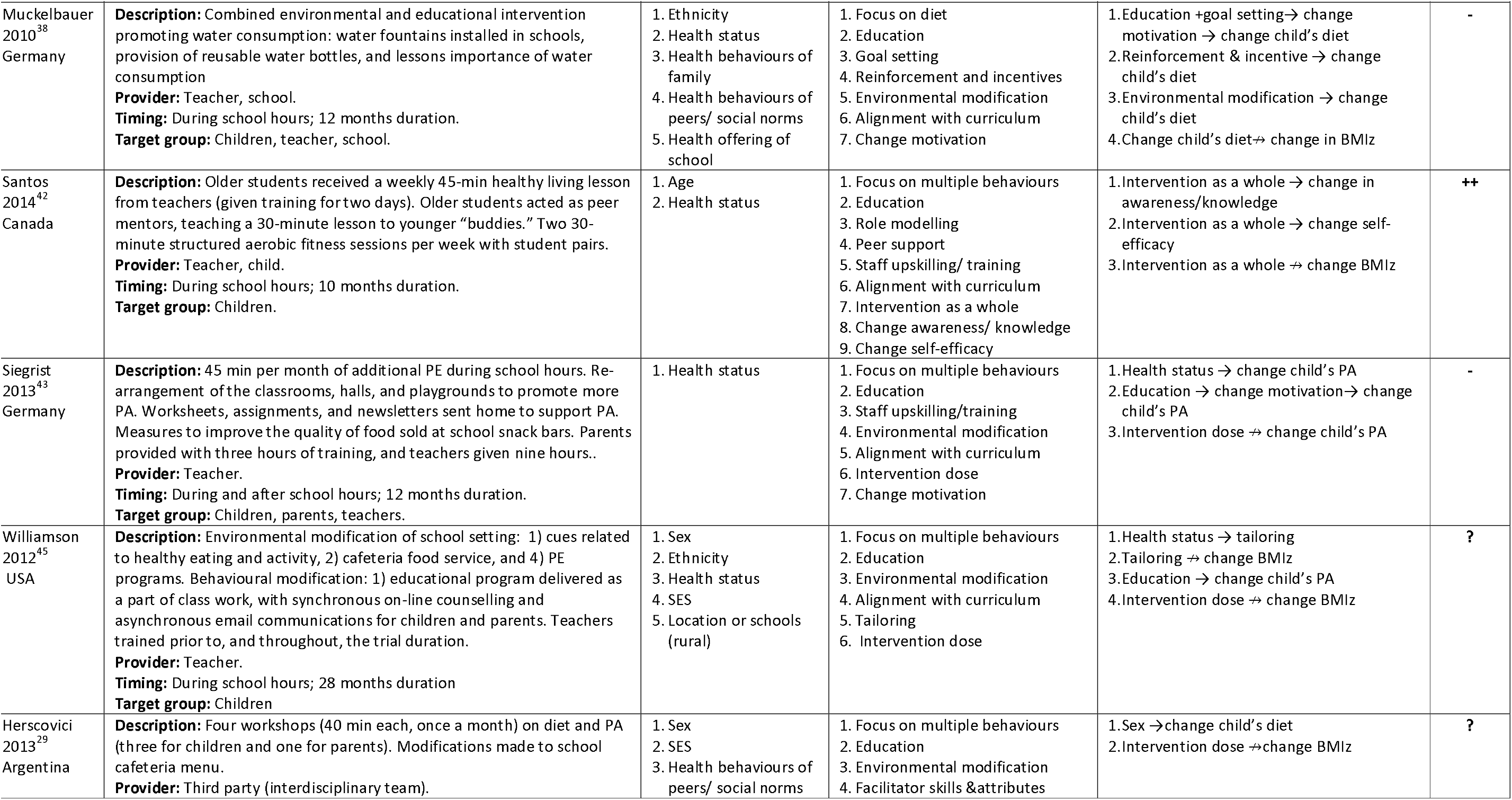
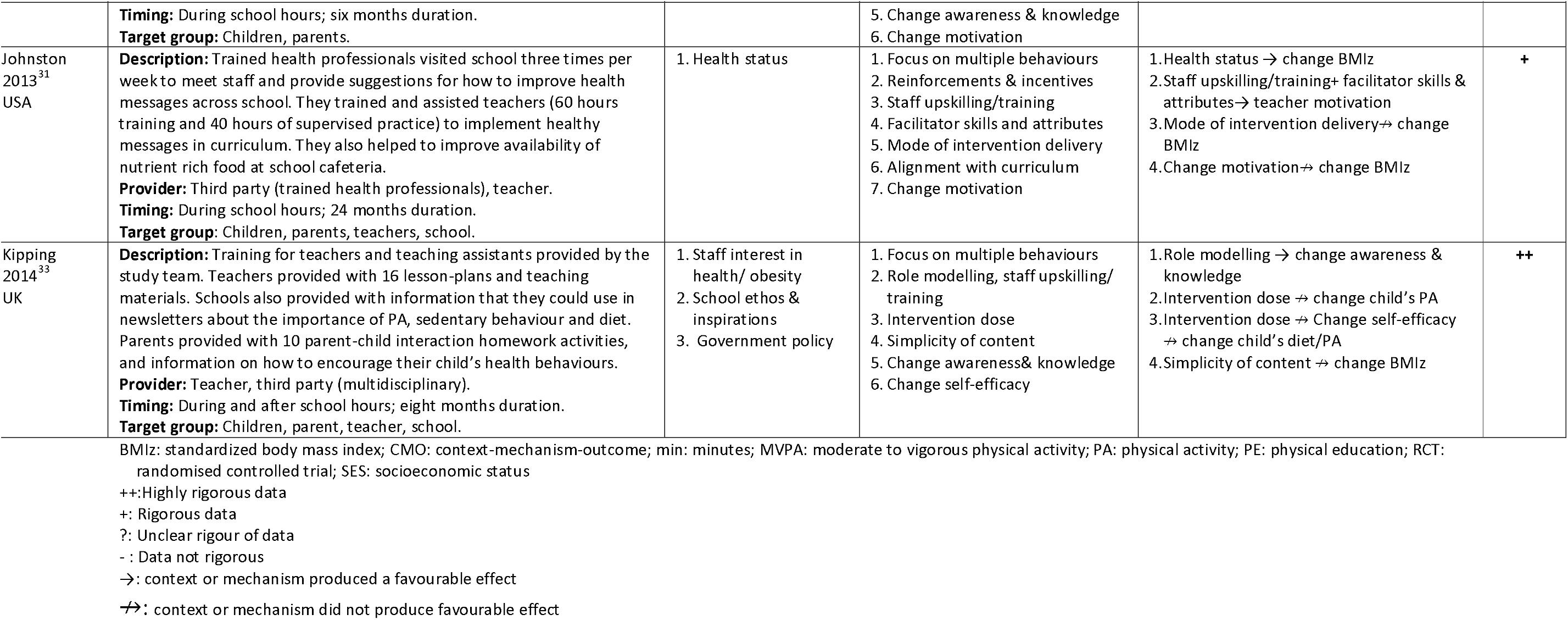
Characteristics of included studies.

The majority of interventions addressed multiple health behaviours (16 studies), followed by diet alone (six studies) and PA alone (three studies). Interventions were most often tested in the USA (six studies), followed by UK and China (three studies in each). Most (n=16) interventions were delivered entirely during school hours and majority interventions (n=13 studies) targeted children, their parents (or family) and teachers together. Teachers were the providers (deliverers) of interventions most often (18 studies) either exclusively (10 studies) or with a third party such as researchers, children, health or PA experts (eight studies). Interventions durations ranged from three months to four years with a median of 12 months (IQR 7.5 to 24).

### 3.3 The final programme theory

Amendments to the programme theory throughout the review period can be seen from figures 1 and 2 and in supporting information section 1. Six new contexts (age, health behaviours of child, pubertal status, parental health status, parental academic attainment, and population health trend) and six new mechanisms (social marketing, timing of intervention delivery, enjoyability of content, simplicity of content, role play, and alignment with curriculum) in total were added to the programme theory over five iterations (available in online supporting information section 1). We found evidence on 16 contexts and 20 mechanisms from the 24 included studies. We present our findings below starting from most cited to least cited contexts and mechanisms across studies.

#### 3.3.1 Contextual factors

**Baseline BMI classification** was a major contextual factor for intervention effect. Four studies found their interventions worked better for children with overweight or obesity in contrast to children of a healthy weight.^22 31 34-36^ Two studies found their intervention worked only for children who were of a healthy weight at baseline.^27 32^ Only one study discretely tailored the intervention differently for the two groups so as to minimize “potential for stigmatizing overweight kids”^45^, albeit with no effect difference in BMIz.

**Sex** appeared to be the next noteworthy context. Girls were reported on several occasions to benefit more from interventions in terms of favourable BMIz, PA or diet change. ^22 25 27 29 34 35^ Study authors argued that girls may be more concerned about their body image and weight, therefore more likely to adhere to the educational content of the interventions. Compared to boys, girls also maintained changes in BMIz after the interventions stopped. ^25^

For **ethnicity**, one study^26^ found evidence that black children benefited more from their intervention than white children. Conversely, another^38^ argued that, since the educational component of the intervention was not tailored to account for cultural differences, their intervention may have been less effective for migrant (non-German) children, although no effect difference by this variable was seen. Two studies ^28,40^ tailored their intervention content for cultural differences and found no difference in the outcomes between children mof different ethnicities.

Older **age** children achieved lower BMIz ^36^ and higher PA levels ^35^ Li et al. ^35^ argued this may be because older children are better able to understand and follow the directions associated with the intervention.

**Parental academic attainment** also impacted an intervention’s effects. In two studies, children of parents with lower academic attainment were less likely to make dietary changes.^24 36^ These children were also less likely to complete the intervention. ^23 24^

**Peer behaviour and social norms** were noted contexts in two studies. ^24 27^ DeRuyter et al. ^24^ who replaced children’s sugary drinks with artificially sweetened ones noted that the social norm among Dutch children to bring a sugar sweetened drink with them to school allowed for easy switch to an artificially sweetened drink. So, the intervention is unlikely to work in countries where sugar-sweetened drinks are not routinely consumed at school. Grydland et al., ^27^ who offered fruit and vegetable snacks at break time noted that fruit, but not vegetable, intake increased amongst the children. They argued that this was because in Norway, vegetables are often eaten during evening meal, which is why only fruit consumption increased.

**Population health trends** appeared to affect how an intervention worked in one study where the population prevalence of childhood overweight and is high, it is unlikely that a simple educational intervention will suffice. ^41^ Other contexts potentially influencing an interventions’ effect on child’s health were **good parental health status**, ^22^ rural **location of school**^40^ and **high socioeconomic status** (SES). ^40^

#### 3.3.2 Mechanisms

**Education** was the most used mechanism (18 studies). Education alone led to a change in motivation in three studies^28 38 43^ and to change in self efficacy in one, ^32^ but not BMIz. Spiegel et al. ^44^ demonstrated that education, when delivered through mechanisms of **goal setting, role play** and **tailoring**, would change knowledge, self-efficacy, and motivation. The knowledge change was argued to have brought about change in child’s diet, PA levels and BMIz. Williamson et al.^45^ provided evidence that education combined with **alignment** with **curriculum** as a mechanism could change child’s PA.

The second most cited mechanism was sufficient **intervention dose**. Three studies argued that sufficient intervention dose brought about a significant BMIz change. ^32 35 36^ Ten ^35^ and 30 ^36^ minutes of integrated daily PA over 12 and 48 months respectively was effective in changing BMIz for children with overweight or obesity. While 70 minutes of intermittent MVPA, five times a week, for nine months was argued as sufficient to change BMIz in children with healthy weight at baseline. ^32^

Several other studies argued that the intervention dose was too low to achieve a BMIz reduction. ^25 27 29 33 37 41 43 45^ However, most of these involved educational health promotions and little enabling of PA. For example, 20 months^27^ and 28 months of PA promotion in school ^45^ was insufficient to alter BMIz compared with the control group. While BMIz stayed unaffected, children’s PA levels improved after three years of 80 min MVPA at least twice a week^28^ but not after 6 months of 10 min daily MVPA^37^. Both interventions claimed to be enjoyable (i.e. an additional mechanism).

Insufficient intervention dose was also proposed as a reason for unchanged diet behaviour ^23^ because the intervention could only influence food consumed within school hours and therefore had limited potential to change total daily intake. Kipping et al. ^33^ hypothesised **self-efficacy** as a mechanism for change in diet and activity behaviours, however suggested that intervention dose was not enough to change self-efficacy. They also suggested that change in PA requires more intense PA interventions, however also noted that given how busy schools and staff already are it may not be feasible.

**Environmental modification** often altered food options available for children but this was not always associated with change in dietary behaviour.^22 23 26 29 40 41 43 45^ Only in two studies^36 38^ was environmental modification associated with a change in child’s diet, and with a BMIz change in one. ^36^ These modifications consisted of: a) modifying the arrangement of school lunches in self-service areas: fruit and vegetables were placed before other options. ^36^ and b) the installation of water fountains in school premises. ^38^ The authors argued that these environmental modifications – once implemented – led to sustainable changes in dietary behaviours.

Two studies used environmental modification as a mechanism to bring about change in the children’s PA levels.^28 43^ Gutin et al. ^28^ created what they termed a “fitogenic environment” through the provision of additional PA afterschool, whilst Siegrist et al. ^43^ made modifications to the classrooms, halls and playgrounds to encourage PA. Both studies demonstrated positive impacts on PA levels, but not on BMIz.

**Intervention as a whole** was cited as a mechanism in six studies. We assume that most interventions are designed to work as a whole, however in the context of this realist review, only a small number of studies were explicit in stating that it was the entirety of the intervention that brought about a change in an outcome, with one of these achieving BMIz change. ^44^ Spiegel et al. ^44^ attributed the BMIz change to the various intervention components (via **role play, goal setting, tailoring, and alignment with curriculum**) working “in concert … creating something greater than the sum of the parts.” Two other studies, Sahota et al. ^41^ and Rosario et al. ^39^, reported that the intervention as a whole only changed dietary intake. Similarly, Foster et al. ^26^ found that their intervention, as a whole, only led to a change in sedentary behaviour, with Grydland et al. ^27^ Rush et al. ^40^ and Santos et al. ^42^ citing their interventions as a whole changed knowledge and awareness of health behaviours.

**Alignment with curriculum and staff upskilling/ training** were often employed together ^22 25 26 31 35 41-43^ aiming to educate the children in order to change the behaviour and yet led to behaviour change in only one study. ^35^ This was achieved via additional contributions from **tailoring** of this intervention to the age group and an optimal intervention dose.

**Tailoring** was employed in four studies ^35 39 44 45^ and, as mentioned above, only in one^35^ it led to the desired behaviour change in children. Tailoring was demonstrated via age-, and space-appropriate exercises where students and teachers were allowed to develop new activities in one study, ^35^ options to increase intensity of aerobic exercises in class in another, ^44^ and a software programme recognizing children with overweight and offering them different content in one. ^45^ The fourth study^39^ ensured that intervention content could be tailored by the teachers themselves in order to best serve the needs of their pupils.

Five studies reported their interventions to have **enjoyable content**. ^28 34 37 39 41^ However, only one of these studies^28^ highlighted that their enjoyable PA content (by offering different activities and enabling children to see their progression) changed motivation.

**Simplicity** of the intervention and / or intervention content was cited in three studies, ^25 30 33^ all from the UK. One argued that their simple message led to change in child’s PA levels. ^25^ The other two studies, ^30 33^ found their simple interventions not successful as a mechanism in changing BMIz. It must be reiterated here that we took authors’ labelling of their intervention as “simple” and there is limited interpretation possible from them. Kipping et al. ^33^ employed child education, role modelling, teacher training, and parent counselling. They argue in their conclusions that such “simple school-based interventions that are designed to minimise costs” cannot bring about major change in diet and PA. Fairclough et al. ^25^ on the other hand although employed education and training for child, teacher, and parent-focused on changing the curriculum to include the simple message ‘move more sit less’ which they believe was a simple non-prescriptive approach.

### 3.4 Gaps in evidence

We found no evidence for some individual contextual factors (such as child’s academic attainment, health literacy, perceived health status and perceived importance of own health), and some family factors (family constraints, family structures and relationships, and household income). Also missing was evidence on type of school (public or private), slack (resource) available in school, staff health status, healthiness of the school environment and curriculum flexibility. The mechanisms not addressed in any studies were monitoring/screening, change marketing/promotion of health offering, and changing social norms.

### 3.5 Reporting of Costs

Eight studies reported cost or resource use (see supporting information section 4). Costs for these varied interventions in current GBP values could range from £12 ^37^ to over £1300 ^32^ per child per year.

### 3.6 Reporting on sustainability of intervention

Eleven studies highlighted intervention features which they believe increased the sustainability of the intervention – see supporting information section 4. These were: stakeholder involvement in intervention design and development, delivering it within the existing resources of the school; collaborating with the relevant authorities and sectors; and adaptable (flexible) intervention content.

### 3.7 Findings of sensitivity analysis

Restricting our analysis to only rigorously conducted studies (n=11; judged either ++ or +), ^23 24 28 30-36 42^ we found the key contexts of influence were still baseline BMI,^31 32 34-36^ parental educational attainment^24 36^ and sex ^34 35^ (see supporting information section 5). Among mechanisms, intervention dose ^23 28 32 33 35^ stood out again along with environmental modification ^23 24 28 36^ as the most often cited.

## 4. Discussion

This realist synthesis found that female sex, and older age, alongside higher parental academic attainment, are key contexts for intervention effectiveness. While some interventions benefited children with a higher baseline BMIz status others benefited already healthy weight children. Girls appeared to benefit from the interventions due to the influence of social norms surrounding body image, which is in line with findings of a recent large scale study in the UK. ^46^ Future studies should therefore consider how interventions may better meet the needs of boys while also addressing the negative social norms surrounding female body image. Similarly, interventions should ensure that they are not just effective for children of highly educated parents, or those without overweight and obesity, because this may inadvertently widen health inequalities.

Despite socio-economic status (SES) being a well-known moderator of intervention effect for health promotion interventions, ^47^ it was formally explored in only one included study. ^40^ This limited evidence on SES was also reported in a recent overview of obesity prevention in adolescents. ^48^ Thus, it is important to consider here how interventions may widen health inequalities if they offer more favourable outcomes for people who are socio-economically better off. As aforementioned, parental education, which is a proxy indicator for SES, ^49 50^ was associated with intervention uptake and effect. Educational attainment is only one domain associated with SES, and so future studies should separate the effects of SES from parental education levels. This will allow us to target the context that is preventing the intended mechanisms from working.

The perceived sufficiency of the intervention dose appeared to affect BMIz in various contexts. However, what constituted sufficient or optimal dose (or dose range) was not specified. Dose can include frequency of and duration of an intervention session (per week or per month) as well as the duration of the entire intervention (in months or years). Which, if any, or what combination of these components may be more beneficial is unknown. A recent systematic review found no link between dose and weight outcomes, which they argued could be either because behaviour change is non-linear or because of varied reporting of dose. ^51^ Given the emphasis placed on intervention dose by many studies in this review, this is a key area for future clarification.

Interventions adopting environmental modification require little individual agency to alter health behaviours, and therefore may be simpler and more sustainable than educational interventions. ^52^ However, the limited evidence on changing BMIz is important as it may suggest further intervention is required to impact health beyond behaviour change. Simplicity and enjoyability of intervention were argued to have the potential to change activity and diet related health behaviours. However, we need clarity on what children deem simple or enjoyable.

Interventions using education as the sole mechanism appeared to have limited impact on behaviour or BMIz. This aligns with the broader evidence base, which suggests educational interventions are unlikely to elicit effective changes for children, ^53 54^ and for general population. ^55^ Relying on individual agency is unlikely to translate into substantial or sustained behaviour change, and consequently obesity prevention. ^56^

### 4.1 Comparison with existing literature

There is no shortage of evidence syntheses of childhood obesity preventive interventions: a recent overview included 66 meta-analyses and systematic reviews on the topic. ^57^ Syntheses usually find that interventions addressing diet and PA are more promising than targeting either behaviour alone. However, the high heterogeneity across studies provided the rationale for our realist synthesis, which aimed to understand the underlying contextual and mechanistic factors that help interventions generate outcomes.

Our findings broadly align with recent realist reviews in the area of childhood PA. ^10 11^ These reviews found that sex (contextual factor) and goal setting, tailoring and intervention dose (mechanistic factors) were linked to the intervention outcomes. Tailoring seldom arose within our review, perhaps due to different operational definitions for what tailoring constitutes or due to the contextual differences between study settings and populations; the review of Hnatiuk et al. ^10^ focused on children aged 0-5 in pre-school settings, whilst the review of Brown et al. ^11^ looked at family-based interventions for children of primary school age (5-12 years). There may be more scope to tailor interventions within these settings in contrast to a primary school setting. While many interventions aimed to align or embed content within the school curriculum, they rarely hypothesised this mechanism to affect BMIz. It may also be that processes were not in place to measure these mechanisms in studies and is not a sign per se that these are ineffective. It would be good in future to consider a-priori how mechanisms would act together to bring about a change and evaluate if the process happened as anticipated.

### 4.2 Strengths and limitations of our realist synthesis

The key strength of this review is that we approached the existing evidence on obesity prevention to understand why and how an intervention works rather than whether it works. The realist synthesis – a relatively new method – allowed us to address these questions which are important to decision-makers. We present new insights into the evidence beyond traditional meta-analysis on the intervention outcome and avenues for future exploration. The findings should help implement an effective obesity prevention intervention in practice.

The review included a large, robust dataset from the most recent Cochrane review. ^8^ We included all qualitative and process evaluations from the 24 studies, amounting to 71 documents in total. This led to rich data for analysing CMO configurations. We restricted our sampling frame to the Cochrane review, which is up to date until 2015 so we may have missed new interventions, contexts, or mechanisms, which is a limitation. The planned Cochrane update effort has identified (but not extracted) a further 162 relevant trials published between 2015 and 2018 and search for trials after 2018 is ongoing. However, the included interventions in the Cochrane review did not change substantially since its first publication in 2002 (i.e., with a downstream focus on individual behaviour change) ^8^ and this was confirmed in a recent secondary analysis of the Cochrane review^58^ using a Wider Determinants of Health lens. The findings indicate that a) the majority of studies target individual dietary and PA behaviours, and b) the focus of childhood obesity prevention interventions has not changed over time since 1993-publication date of the oldest study included in the Cochrane review.

This is a limitation of the evidence-base, whereby the focus is traditionally on behaviour change at individual levels, and environmental or policy interventions targeting wider determinants of health (upstream) are rarely evaluated in randomised trials. ^7 48^ Policy interventions can be evaluated using randomised designs, ^59^ where one geographical or political region may implement the policy sooner than others (waitlist control or stepped wedge design). Where randomisation is not feasible, interrupted time series or controlled before after designs could be employed to evaluate wider determinants of health and policy interventions. ^60^ That said, two recent systematic reviews^7 61^ of natural experiment studies also found that the included studies predominantly focused on downstream determinants of childhood overweight and obesity. Thus, we anticipate it is unlikely that the focus of interventions has changed dramatically between 2015 and 2020.

### 4.3 Implications for UK-based primary schools

The stakeholder consultation indicated that UK primary schools have limited resources to take on obesity prevention tasks. With no evidence in the review to support usefulness of additional health education for changing BMIz, it may be difficult to justify teachers doing this. Education may be important but is insufficient on its own to change BMIz. Implementing environmental modification (such as the installation of water fountains, changed canteen offerings) may be perceived more favourably by school staff. This may also bypass the reliance on individual agency for behaviour change. One suggestion^62^ to optimise implementation of a school intervention is to involve delivery staff (school staff, management or third party) in the design and development of the intervention. We recommend including children in this planning.

Given the limitation of school finances in the UK, cost is a major consideration for any intervention. Whilst obesity prevention interventions are likely to be cost effective in the long-term, ^63^ these returns may not be seen by the education sector (or individual schools), and thus the immediate investment required to establish a new initiative may be negatively perceived by stakeholders. Unfortunately, there was insufficient information in the studies to analyse the costs of different intervention types. We need full cost reporting for future interventions, including a breakdown of the costs per intervention component, to facilitate decision making.

## 5. Conclusions

Our findings indicate that being female and older, and having parents with a high academic attainment can help children benefit from obesity preventive interventions, while baseline BMI can affect intervention outcomes variably. The potential ramifications for health inequalities with these contexts must be kept in mind by both commissioners and researchers. Sufficient intervention dose and environmental modifications in schools are mechanisms that may help achieve desired outcomes. In addition, an intervention that worked as a whole rather than a collection of separate components can better achieve desired outcome, illustrating the interdependent nature of the intervention mechanics - the effect being greater than the sum of its parts. That said, few mechanisms favourably influenced BMIz, and were more likely to only change knowledge, motivation, and dietary and physical activity behaviours.

## Data Availability

The data that support the findings of this study are already available in the public domain.

## Contribution of Authors

SI contributed to all stages of the review and wrote the manuscript; JN contributed to all stages of the review and edited the manuscript; LJ and RJ advised as subject experts at all stages and edited the manuscript; TM contributed to data collection and edited the manuscript; JS advised as method expert on rigour assessment and edited the manuscript.

## Acknowledgements

We are grateful to Dr Jill Hnatiuk who provided expert methodological advice.

