## supporting information for "Preventing childhood obesity primary schools: a realist review from UK perspective"

#### Section 1 programme theory development

**V2: Adapted based upon stakeholder survey and initial piloting**

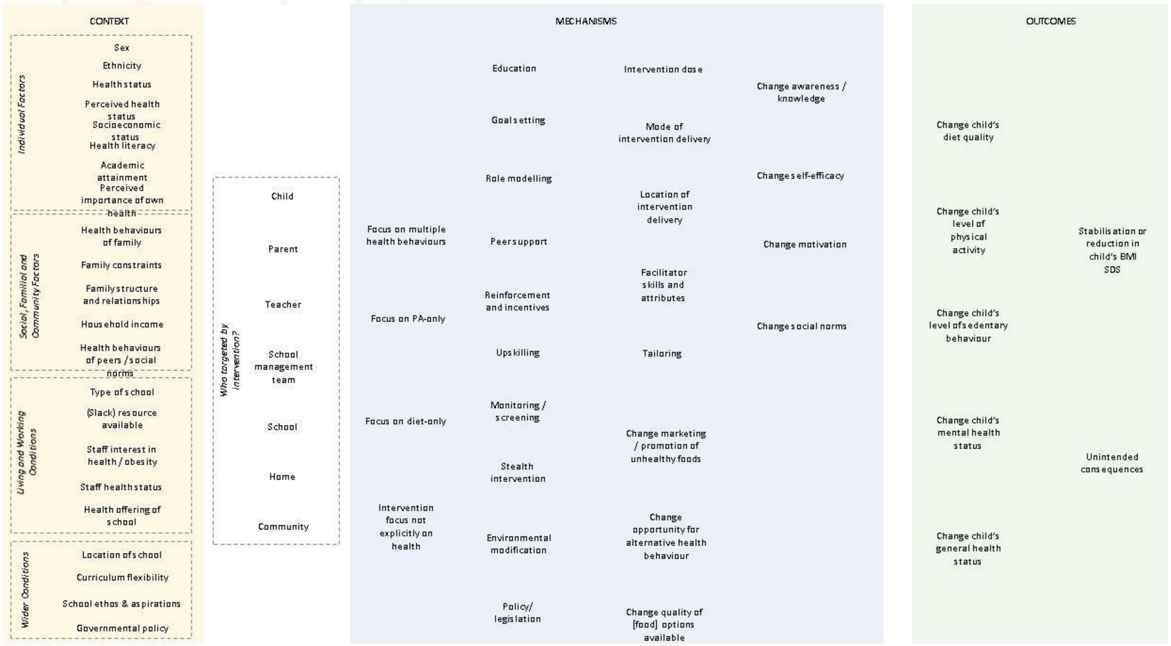

**V3: Refinement after further testing**

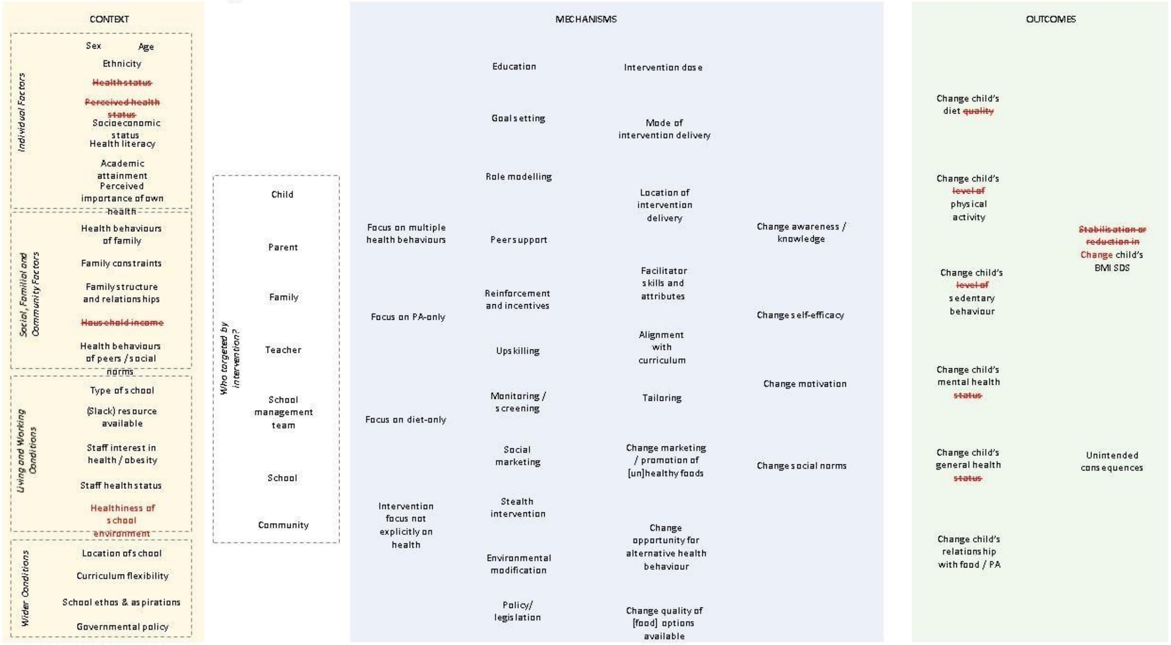

**V4: Refinement during data extraction**

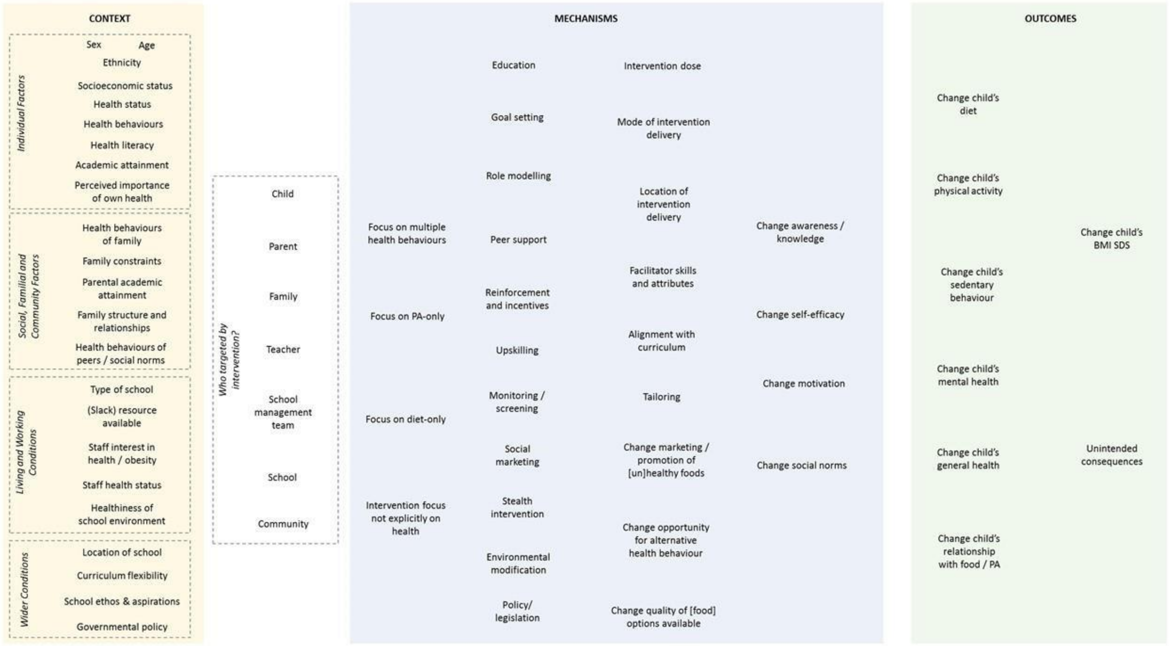

**V5: Further refinement during data extraction**

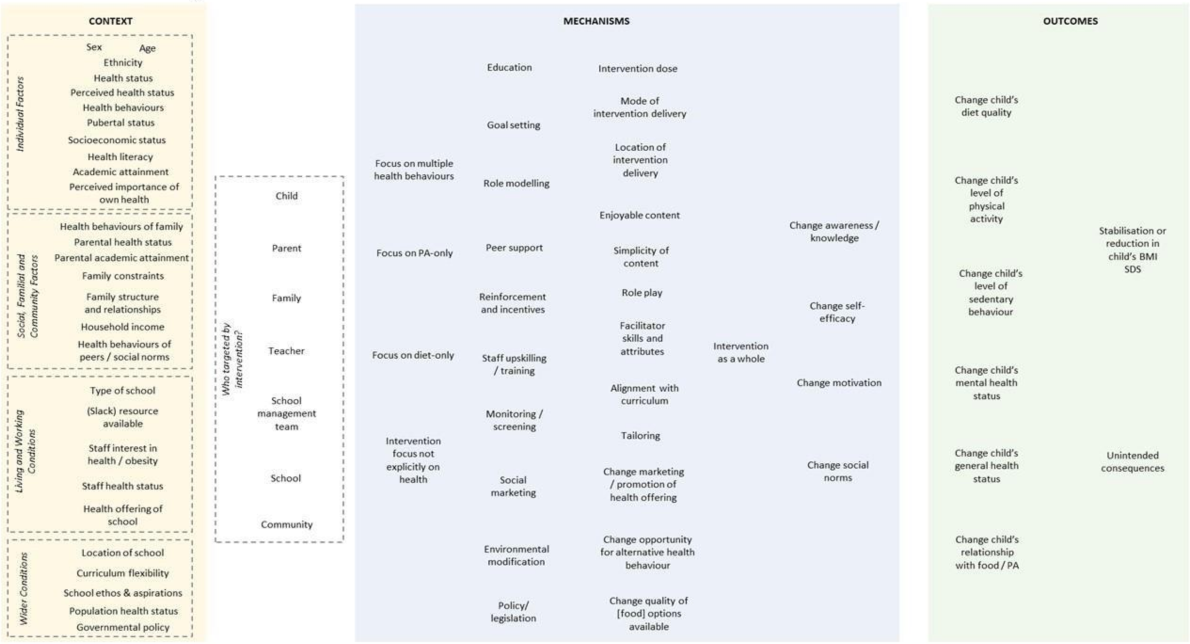

V6: Final

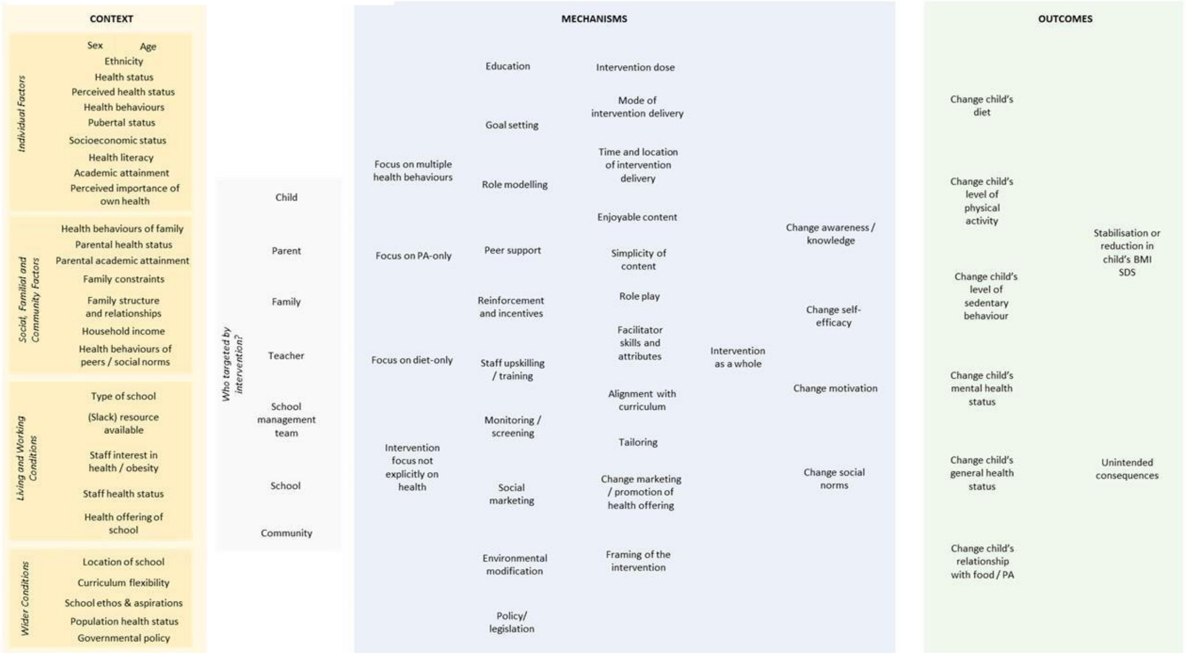
CONTEXT
Individual Factors
Sex
Age
Ethnicity
Health status
Perceived health status
Health behaviours
Pubertal status
Socioeconomic status
Health literacy
Academic attainment
Perceived importance of own health
Social, Familial and Community Factors
Health behaviours of family
Parental health status
Parental academic attainment
Family constraints
Family structure and relationships
Household income
Health behaviours of peers / social norms
Living and Working Conditions
Type of school
(Slack) resource available
Staff interest in health / obesity
Staff health status
Health offering of school
Wider Conditions
Location of school
Curriculum flexibility
School ethos & aspirations
Population health status
Governmental policy
Who targeted by intervention?
Child
Parent
Family
Teacher
School management team
School
Community
MECHANISMS
Focus on multiple health behaviours
Focus on PA-only
Focus on diet-only
Intervention focus not explicitly on health
Education
Goal setting
Role modelling
Peer support
Reinforcement and incentives
Staff upskilling / training
Monitoring / screening
Social marketing
Environmental modification
Policy/ legislation
Intervention dose
Mode of intervention delivery
Time and location of intervention delivery
Enjoyable content
Simplicity of content
Role play
Facilitator skills and attributes
Alignment with curriculum
Tailoring
Change marketing / promotion of health offering
Framing of the intervention
Intervention as a whole
Change awareness / knowledge
Change self-efficacy
Change motivation
Change social norms
OUTCOMES
Change child's diet
Change child's level of physical activity
Change child's level of sedentary behaviour
Change child's mental health status
Change child's general health status
Change child's relationship with food / PA
Stabilisation or reduction in child's BMI SDS
Unintended consequences

### Section 2 data extraction template

| DE item | Comments | Text copied in |
|---|---|---|
| First author and date of publication of document(s) mark which paper the data came from below (e.g. (James 2004) or (James 2007)) | | |
| Study location city (country), add text where it is of relevance to context | | |
| Study design(s) (RCT, Mixed method, qualitative, etc.), add text if they justify the methods | | |
| Duration of intervention (of study) in months | | |
| Duration of follow up (for outcome assessment) in months | | |
| Timing (during/ before/ after school) | | |
| Aim (briefly in authors' own words if possible) | | |
| Incentive provided for participation: Monetary or other(s) | | |
| Cost of intervention reported (convert to GBP 2019 value) | | |
| Context: list all reported by matching with the program theory diagram - then also add any additional ones they refer to that are not in our theory diagram but should be<br><br>(N = Number of contexts identified) | | |
| Intervention characteristics (target group, providers, time, duration, intensity, components in authors' words- copy text in) | | |
| Mechanisms triggered-match with program theory diagram and list all that fit- then also add any additional ones they refer to that are not in our theory diagram but should be<br><br>(N= Number of mechanisms identified) | | |
| Outcomes – keep this simple and list only those matched with the program theory diagram | | |

| |
|---|
| (N =Number of outcomes identified) |
| Conclusions of the authors in their words |
| Relevance: data (text sections) within a study that show relevance to our theory (for either development or testing), |
| Rigour: study's methods of data collection and analysis and whether we can trust what their claims are |

#### Section 3 example of data extraction

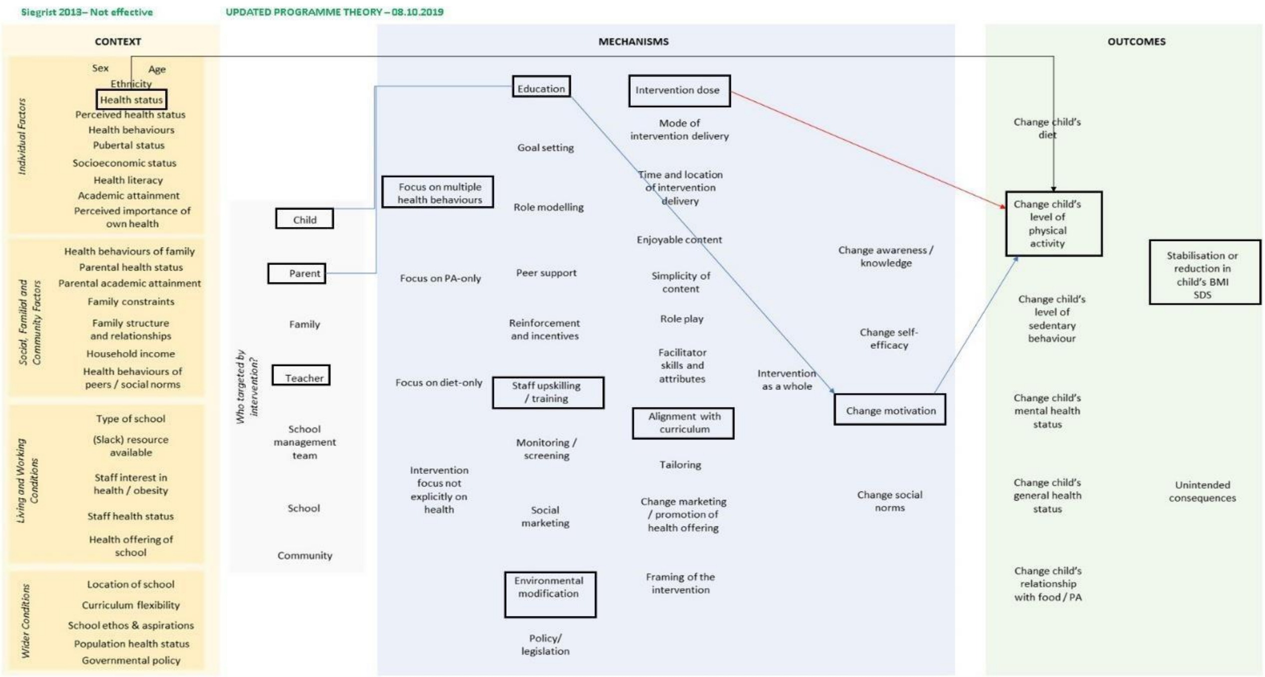

**Context**:

**Health status**-overweight kids had improved PA after intervention: Overweight and obese children showed lower baseline fitness levels. After 1 year, they had significantly improved in two test items (goal throwing and jump and reach) and slightly increased their total score in contrast to a decrease in the controls. *Siegrist 2013 Page 329 para 2* **Mechanisms:**

**Education /aligned with curriculum/ environmentmodification/ multiple behaviours:** The focus of the multifaceted JuvenTUMintervention was on directly educating and encouraging children, teachers, and parents to live active and healthy lifestyles. Additionally, school environmental settings (e.g. the physical environment, organization of school breaks, playing during school time, and sports facilities) were altered to promote more physical activity. These changes were designed to increase physical movement, promote healthier food availability and choices (more vegetables and fruits and less energy-dense food), and reduce media consumption (for further details about the program, please visit our website http://www.juventum.med.tum.de). *Siegrist 2013-page 325 para 2-3*

**Dose (duration) not enough for PA change** Although physical activity remained unchanged in the control group, no significant difference between both groups was found after 1 year. It is possible that the sensitivity of physical activity questionnaires is inadequate to allow for the detection of significant group differences or that the intervention time was to short (10 lessons over a period of 1 year). These questions are being investigated in an ongoing longer term intervention. *Siegrist 2013 page 328 para 4* **.**
*That's not reliable data then is it? don't buy this argument though…because they have used valid PA measure: https://www.ncbi.nlm.nih.gov/pubmed/11343497*
**environment modification> change in quality of food / changed opportunity for alt health behaviour:** Additionally, measures were taken to improve the quality of food sold at school snack bars and school stores as well as to arrange the classrooms, halls, and playgrounds in a way to promote more physical activity. All teachers of the intervention schools (IS) took part in these trainings.*Siegrist2013-page 325 para 3-6*
**Education > increased motivation and competence? Change in PA:** Successful learning experiences in skill development are particularly important in overweight children to enhance children's perception of competence and motivation, which in turn maximizes participation in physical activity.*Siegrist2013 Page 329 para 2*
**Teacher training/ upskilling:** Three teacher trainings (9 h total) were conducted with the objective of increasing their students' physical activity during lessons and breaks and improving physical education within their schools.*Siegrist 2013-page 325 para 6*
**Parents targeted with education:** Parents participated in two training sessions in which they were given a program overview and practical instruction about health issues (3 h total). They were informed about the development and course of the intervention program, received health-related journals, participated in practical instruction based on increasing motivation to spend more time being active with their children, and were asked to improve health behaviors(e.g. making healthy food choices and less media consumption) with their family. 30% participated.*Siegrist 2013-page 325 para 5*
***Rigour comment:*** issues with their arguments-see above. Plus, they say they lack control for dietary behaviours in the study. But they added info on diet in education as part of the intervention – if not then why add it? Randomisation was unclear; rest of the RoB items were at low risk and analysis appropriate.

#### Section 4 cost and sustainability data

| Study and country | Cost information | 2020 cost in GBP (https://eppi.ioe.ac.uk/costconversion/default.aspx) |
|---|---|---|
| Marcus 2009 Sweden | The intervention was aimed to be financed within the resources of the ordinary school budget. | NA |

| | | |
|---|---|---|
| Khan 2014 USA | Funding figure for 2008 on NIH website states $341,654 for the school year. | £1316.71 per child / year Assuming this is the total cost and divide by number of participants (220) = $1553 per Child per year. |
| Kipping 2014 UK | The main cost drivers for the intervention were the claims by the schools for replacement teachers needed to cover the teachers' attendance at the training days (£5.00 per pupil, £5095.00 in total); the trainers' fees (£2.00 per pupil, £2166.00 in total); the time spent by the research staff organising and attending the training days (£2.00 per pupil, £2492.00 in total); and the printing costs of the materials for the AFLY5 lessons and the homework (£6.00 per pupil, £6694.00 in total). We estimated the opportunity cost of implementing AFLY5 in schools (i.e. the cost of teaching AFLY5 minus the cost of teaching the usual curriculum based on data from schools in the control arm) to be £0.05 per pupil. The costs varied by school, ranging from approximately £13.00 to £36.00 per pupil. The variations between the schools were driven by the costs of the teachers' attendance at the training days. The cost–consequence analysis showed that, for the three secondary outcomes that were affected by the intervention, it cost £18 per child (£18,944 in total) to reduce self-reported time spent on screen viewing at the weekend by 20.86 minutes, self-reported consumption of snacks by 0.22 snacks per day and self-reported consumption of high-energy drinks by 0.26 servings per day. From the teachers' perspective, teachers spent, on average, more time travelling to the training day venue than they usually spent travelling to school; this equated to an additional 0.68 minutes' travel time per pupil, generating an extra cost of £0.19 per child (£206 in total). *page 43 of PHR full report 2016* Results of the economic analysis showed that the cost per child from a school and provider perspective of implementing the intervention was £18 per pupil (£18,944 in total). *PHR full report discussion page 64 para 3* | £20.03 per child/ 8 month |
| Damsgaard 2014 Denmark | NR.<br>**Comment: They say in protocol they measured costs but these are not reported in any of the outcome papers** | NA |

| | | |
|---|---|---|
| Meng 2013 China | social perspective- For year 2013-all costs reported in table 3 and 4; ICERS in table 5<br>The intervention costs per child in combined intervention group was RMB182.4 ($26.8), which was 2.4 times higher than that in the nutrition intervention (RMB52.8, $7.8) or in the PA intervention (RMB52.3, $7.7). page 5 para 3<br>conversion to GBP 2019: costs per child in combined intervention group= £24.1 cost per child nutrition intervention= £ 7.04 cost per child PA intervention= £ 6.97 | Combined intervention: £21.05 per child/ 6 month Diet intervention: £6.13 per child/ 6 month PA intervention: £6.05 per child/ 6 month |
| Mucklebauer 2010 Germany | In our study, the initial costs per water fountain were ~2500 euros and the long-term costs per enrolled child were _13 euros per year. The educational intervention was presented by the teachers; therefore, no additive costs emerged.<br>Muckelbauer 2009—discussion)<br>The schools were provided the running costs of the water fountains (about 800 Euros/year) by the study budget in the intervention period and the following year and had to pay the costs themselves afterwards. Muckelbauer 2009a page 852- [para 8)<br>11 out of 17 schools succeeded in keeping the fountains (Table 1). Headmasters of the schools that kept the fountains reported that maintaining costs were paid by fees from parents (*n* = 6), bounties (*n* = 3), school fund (*n* = 4), and other school-related associations (*n* = 3), solely or in combination. (Muckelbauer 2009a page 853- para 5)<br>**Comment: These are values for year 2007.**<br>**Unclear how educational intervention had 'no additive' cost- at least materials would have a cost.** | Cost of fountain: £2643.85<br>Cost of yearly maintenance: £846.03<br>Cost for 2 years: £3,489.88<br><br>For 2500 children the cost per child for first 2 years: £1.39<br><br>For 250 children the cost per child for first 2 years: £13.9 |
| | **Long term not defined**<br>**Cost of bottles not reported** | |
| Rush 2012 NewZealand | The programme is cost-effective, the main costs are the salaries of the Energizers and team leader and the travel required to move between schools. We calculate that the average cost of the intervention for each child, each year, is less than $40 New Zealand and this could be improved by further efficiencies. *Rush 2012 page 585 para 1* | £22 per child/ year |
| Grydeland 2014 Norway | NR.<br>**Comment: In protocol they say they were collecting cost data for CE analysis and in the final paper they say it didn't cost much as teachers were the main providers.** | NA |
| Paineau 2008 | Family dietary coaching has an individual cost of around 1 €/d/person (US $1.42/d/person), which should be compared with the cost implications of obesity for health care and society.<br>**Comment: Unclear if it is their cost or taken from somewhere else.** | NA |

| Study and country | Supporting text from study | Key facilitators |
|---|---|---|

| | | |
|---|---|---|
| Flexibility considerations | | |
| Li (2010)<br>China | *Exercise intensity varied by grade levels because of the different activities involved and the different rhythm and extent of the same activity. Students in lower grades were more active than those in higher grades. Students in grade 3 were most active, with the highest energy expenditure and intensity of a Happy 10 session.[…] The program provided a variety of safe, moderate, age-, and space-appropriate exercises . Teaching materials included activity cards, video demonstrations, tracking posters, and stickers. Each activity card introduced one exercise and explained how to perform it. The videos showed students from the pilot study performing the activities. Teachers could either demonstrate the activity or show it on a video. The tracking poster and stickers were used to illustrate the progress of each class. <u>Students, teachers and parents were encouraged to develop new activity models, so did the program staffs</u>. Many new programs, much more than that directly from TAKE 10!, were developed, such as<br>"Story in zoo"; "story in farm"; "who is wearing yellow today"; "time like a colt"; "happy and health"; "little frog". –* <u>pg 181</u> | 1. Provide teachers with several options for how to deliver elements of the intervention.<br>2. Teachers able to adapt the intervention based on their knowledge of the pupils ability and preferences.<br>3. Co-produce materials with children, families and teachers (for sustainability). |
| Rosario (2012)<br>Portugal | *Our approach was to standardize recommendations to teachers, allowing them enough flexibility to create interactive interventions and pedagogic instruments to be used with children. This is contrary to previous school-based interventions that have used tight controls to ensure uniform implementation but required frequent staff training and ongoing supports. –* <u>pg 1364</u> | 1. Ensure recommendations for teachers are consistent, however allow flexibility in how the information is delivered. |
| Sustainability considerations | | |
| Marcus (2009)<br>Sweden | *The programme was designed to be an integrated, sustainable part of the ordinary school curriculum, possible to maintain within the ordinary school budget. –* <u>pg. 415</u> | 1. Embedding the intervention within the curriculum. |
| Muckelbauer (2009)<br>Germany | *The schools were provided the running costs of the water fountains (about 800 Euros/year) by the study budget in the intervention period and the following year and had to pay the costs themselves afterwards. –* <u>pg. 852</u><br>*11 out of 17 schools succeeded in keeping the fountains (Table 1). Headmasters of the schools that kept the fountains reported that maintaining costs were paid by fees from parents (n = 6), bounties (n = 3), school fund (n = 4), and other school-related associations (n = 3), solely or in combination. -* <u>pg. 853</u> *A widespread transfer of our programme is favoured by the fact that implementation was exclusively based on school staff. This independency of external support enhances the practicability and sustainability of the programme. However, external financial support may increase the number of schools maintaining the fountains when the initial installation of the fountains has been* | 1. Ensure educational components can be delivered by school staff.<br>2. Target intervention at school staff rather than children. Reduces need for external support.<br>3. Ensure financial cost of maintaining intervention is low and available (€800). |
| | *managed. The fact that even in deprived districts, the vast majority of schools were willing and able to find a financing for the fountains is of particular significance –* <u>pg 856</u> | |
| Kipping (2014)<br>UK | *Although the quantity of lessons and homework assignments delivered was high, the difficulties of incorporating some of the AFLY5 materials into more technologically advanced and interactive current teaching practice, coupled with pressure on teachers' time and a need to adapt the materials to suit students' differing abilities and ensure their engagement, resulted in mixed enthusiasm for AFLY5.–* <u>pg. 70</u> | 1. Ensure intervention resources are delivered in a similar format to existing school resources.<br>2. Ensure resources do not require much modification by teachers. |
| Gutin (2008)<br>USA | *Institutionalization of innovative programs requires that the program be built on existing infrastructure and resources. MCG FitKid was built on the infrastructure in the schools (i.e., school teachers and paraprofessionals, facilities, and transportation system), which increases the potential for success of the program and the likelihood of adoption in other schools and communities. -* <u>Yin</u> <u>2005, pg. 2160</u> | 1. Limit the need for additional infrastructure and resource when implementing a school-based intervention. |

| | | |
|---|---|---|
| Grydeland (2014)<br>Norway | *The results of this intervention study are important to public health, as feasibility and sustainability were high priorities when designing the intervention. This has been recommended in previous studies and reviews [6,8,37]. Although comprehensive, the intervention components were designed to be able to fit into current school curricula without substantial extra costs. With limited instructions and material provided by the study group, teachers were key deliverers of the intervention components. No extra personnel or costly material are needed to carry out such components in the current school system, and all components could easily be incorporated into existing curricula for this age group. –* <u>pg. 11</u> | 1. Align intervention content with the school curriculum.<br>2. Ensure intervention can be implemented for limited additional cost.<br>3. Intervention content should be simple and easy for teachers to deliver. |
| Fairclough (2013)<br>UK | *Interventions that can be implemented by school personnel in 'real life' conditions (i.e., without re searcher support and resources) are advocated [15], as these are less costly [13], and are more likely to be integrated within existing curricula and sustained over time. –* <u>pg. 2</u> | 1. Ensure intervention can be delivered without additional resource from study team.<br>2. Align intervention content within the curriculum. |
| Stakeholder involvement considerations | | |
| Foster (2008)<br>USA | *Each school formed a Nutrition Advisory Group to guide the assessment. Teams included administrators, teachers, nurses, coaches, and parents. After completing ratings on healthy eating and physical activity, schools developed an action plan for change. –* <u>pg. 795</u> | 1. Create diverse stakeholder advisory groups and include in intervention design and delivery. |
| Fairclough (2013)<br>UK | *The intervention design and content were informed by formative work conducted with parents, children, and teachers in 10 of the schools in the year prior to intervention commencement [22,23]. –* <u>pg. 3</u> | 1. Design intervention with the input of target recipients and delivery staff. |
| Cao (2015)<br>China | *One important feature of this intervention model was the collaboration between the Education Bureau and Institute of Education, which guaranteed the sustainability of the intervention.* <u>pg. 554</u> | 1. Consider collaborating with government departments.<br>2. |

### Section 5 additional analyses

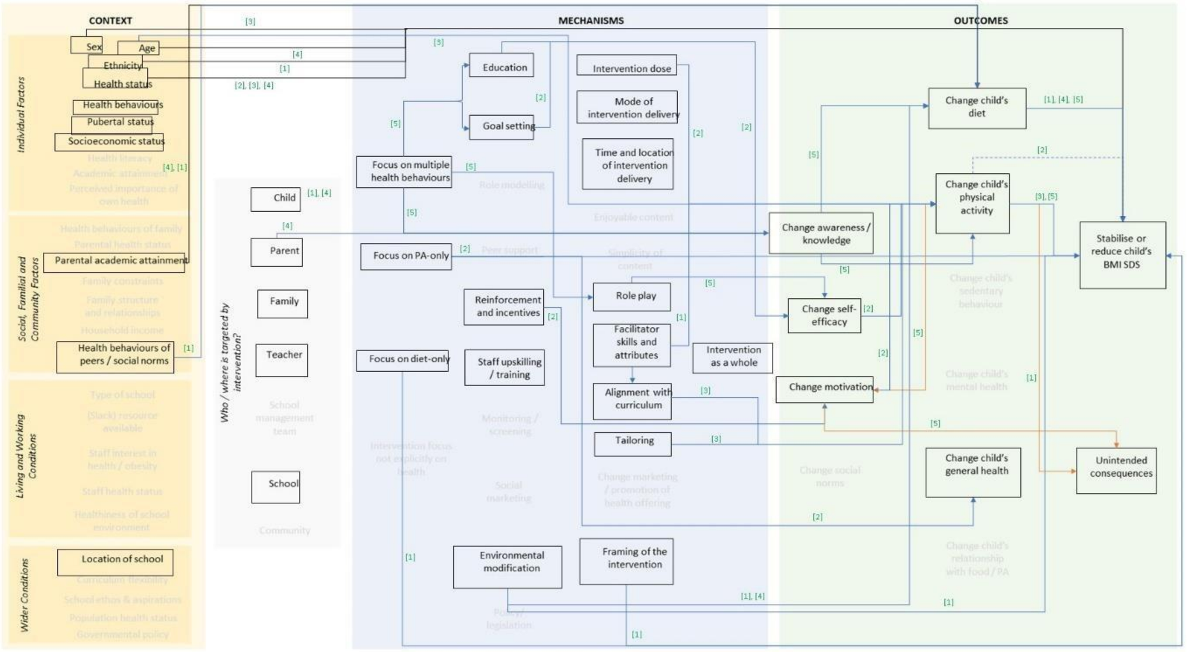

*Figure 1synthesis of effective studies*

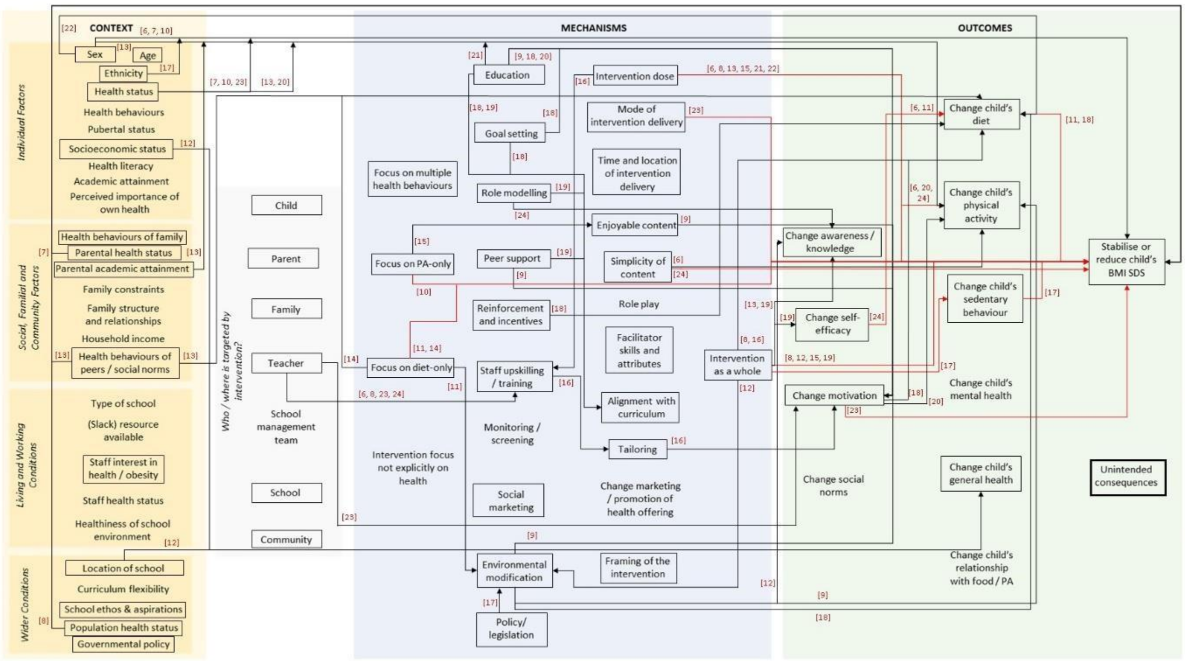

*Figure 2 synthesis of ineffective studies*

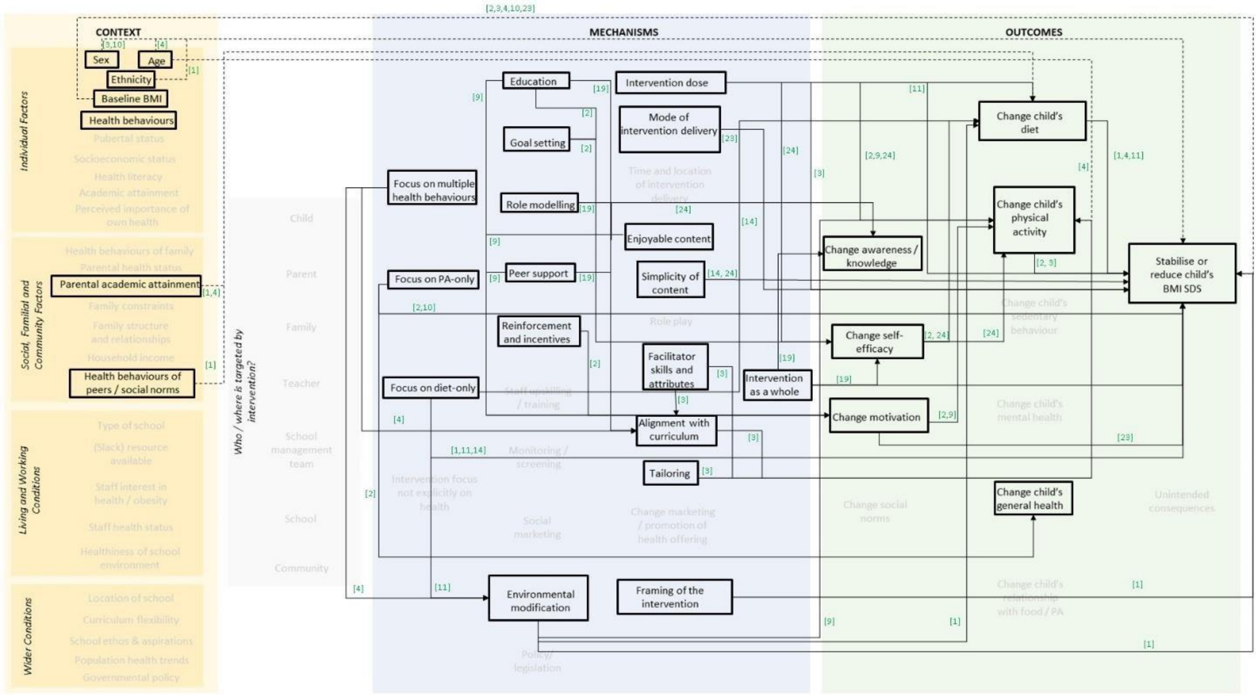

*Figure 3- analysis of rigorous studies*

#### Section 6

details of study flow

**PRISMA 2009 Flow Diagram**

**Prevention of obesity in primary school children: Realist review**

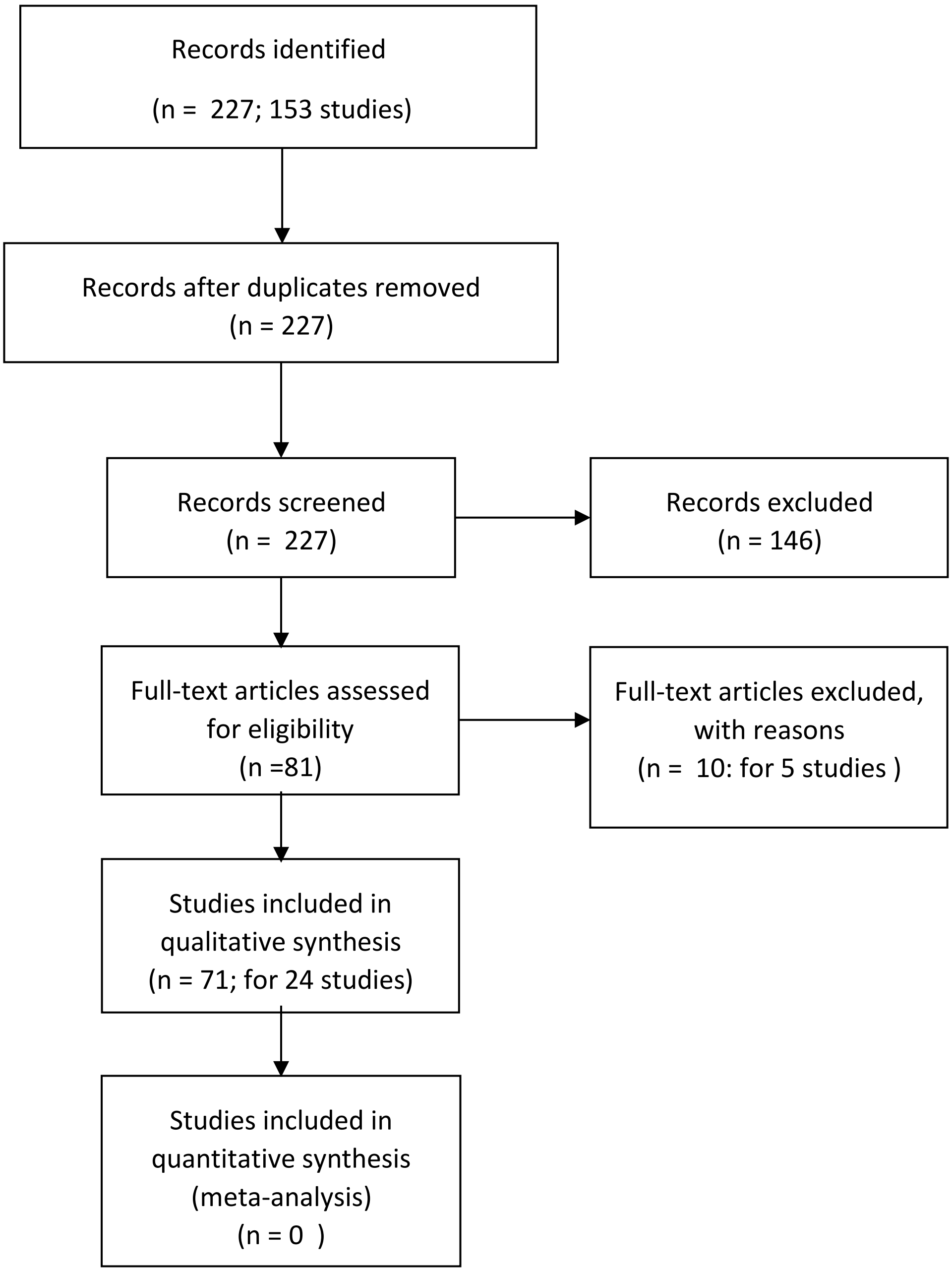

#### Excluded study table

| Study ID | Reason for exclusion | Reference |
|---|---|---|
| Haire-Joshu 2010 | The intervention setting is outside of school and class and in community only | Haire-Joshu, D. N., M. S. Elliott, M. Davey, C. Caito, N. Loman, D. Brownson, R. (2010). "The use of mentoring programs to improve energy balance behaviors in high-risk children." Obesity **18 Suppl 1**: S75-S83. |
| Robinson 2010 | The intervention setting is outside of school and class and in community only; | Robinson TN, Matheson DM, Kraemer HC, et al. A randomized controlled trial of culturally tailored dance and reducing screen time to prevent weight gain in low-income African American girls: Stanford GEMS. Arch Pediatr Adolesc Med. 2010;164(11):995–1004. doi:10.1001/archpediatrics.2010.197 |
| HEALTHY STUDYGP 2010 | Intervention setting is middle school | HEALTHY Study Group. A school-based intervention for diabetes risk reduction. New England journal of medicine 2010;363(5):443-53<br>Hall WJ. School factors as barriers to and facilitators of a preventive intervention for pediatric type 2 diabetes. Translational Behavioral Medicine 2014;4(2):131-40.<br>Marcus MD, Foster GD, Ghormli L. Shifts in BMI category and associated cardiometabolic risk: prospective results from HEALTHY study. Pediatrics 2012;129(4):e983-91.<br>Volpe SL, Hall WJ, Steckler A, Schneider M, Thompson D, Mobley C, et al. Process evaluation results from the HEALTHY nutrition intervention to modify the total school food environment. Health Education Research 2013;28(6):970-8.<br>Schneider M, Hall WJ, Hernandez AE, et al. Rationale, design and methods for process evaluation in the HEALTHY study. International Journal of Obesity (2005). 2009 Aug;33 Suppl 4:S607. DOI: 10.1038/ijo.2009.118.<br>HEALTHY Study Group. HEALTHY study rationale, design and methods: moderating risk of type 2 diabetes in multi-ethnic middle school students.Int J Obes (Lond). 2009 Aug;33 Suppl 4:S4-20. doi: 10.1038/ijo.2009.112. |
| Amaro 2006 | Intervention setting is middle school | Amaro, S., A. Viggiano, A. Di Costanzo, I. Madeo, A. Viggiano, M. E. Baccari, E. Marchitelli, M. Raia, E. Viggiano, S. Deepak, M. Monda and B. De Luca (2006). "Kaledo, a new educational board-game, gives nutritional rudiments and encourages healthy eating in children: a pilot cluster randomized trial." Eur J Pediatr **165**(9): 630-635. |
| Paineau 2008 | Intervention intervention setting is outside of school /class and in community only | Paineau, D. L., F. Beaufils, A. Boulier and et al. (2008). "Family dietary coaching to improve nutritional intakes and body weight control: A randomized controlled trial." Archives of Pediatrics & Adolescent Medicine **162**(1): 34-43. |

#### Included study documents

**Study ID,  Location (Refs)**

1. de Ruyter 2012, Netherlands (1-6)
2. Khan 2014 ,USA (7-9)
3. Li 2010, China (10, 11)
4. Marcus 2009, Sweden, (12)
5. Spiegel 2006,USA,(13)
6. Fairclough 2013, UK, (14-17)
7. Cao 2015 China,(18, 19)
8. Sahota 2001 ,UK, (20, 21)
9. Gutin 2008 USA, (22-24)
10. Lazaar 2007 France, (25)
11. Damsgaard 2014,Denmark ,(26-32)
12. Rush 2012 New Zealand, (33-35)
13. Grydeland 2014 Norway,(36-40)
14. James 2004 ,UK, (41, 42)
15. Meng 2013 China,(43, 44)
16. Rosario 2012 Portugal,(45-47)
17. Foster 2008 , USA,(48)
18. Muckelbauer 2010 Germany,(49-53)
19. Santos 2014 Canada, (54, 55)
20. Siegrist 2013 Germany, (56-58)
21. Williamson 2012 USA, (59-62)
22. Herscovici 2013 Argentina, (63)
23. Johnston 2013,USA,(64)
24. Kipping 2014, UK,(65-71)

**References  to the 71 documents**

1. de Ruyter JC. The effect of sugar ‐ free versus sugar ‐ sweetened beverages on satiety, liking and wanting: An 18 month randomized double ‐ blind trial in children. Study Protocol and amendments. 2013.

2. de Ruyter JC, Katan MB, Kuijper LD, Liem DG, Olthof MR. The effect of sugar-free versus sugar-sweetened beverages on satiety, liking and wanting: an 18 month randomized double-blind trial in children. PLoS One. 2013;8(10):e78039.

3. de Ruyter JC, Olthof MR, Kuijper LD, Katan MB. Effect of sugar-sweetened beverages on body weight in children: design and baseline characteristics of the Double-blind, Randomized INtervention study in Kids. Contemp Clin Trials. 2012;33(1):247-57.

4. de Ruyter JC, Olthof MR, Seidell JC, Katan MB. A trial of sugar-free or sugar-sweetened beverages and body weight in children. N Engl J Med. 2012;367(15):1397-406.

5. Katan MB, de Ruyter JC, Kuijper LD, Chow CC, Hall KD, Olthof MR. Impact of Masked Replacement of Sugar-Sweetened with Sugar-Free Beverages on Body Weight Increases with Initial BMI: Secondary Analysis of Data from an 18 Month Double-Blind Trial in Children. PLoS One. 2016;11(7):e0159771.
6. Olthof MR. A Study of the Effect of Replacing Sugary Drinks by Low-sugar Alternatives on Body Weight and Fat Mass in Children (DRINK). 2009.

7. Hillman CH. ERPS to academics: exercise effects on  cognition in school-aged children. Research Portfolio Online Reporting Tools. [Project Funding Information for 2008]. In press 2008.
8. Hillman CH. Exercise Effects on Cognition in School-Aged Children (FITKids). 2011.

15. Fairclough SJ. The CHANGE! (Children's Health, Activity and Nutrition: Get Educated!) Project: a clustered randomised controlled trial. 2011.

33. Graham D. Project Energize. 2010.

40. Lien N, Bjelland M, Bergh IH, Grydeland M, Anderssen SA, Ommundsen Y, et al. Design of a 20-month comprehensive, multicomponent school-based randomised trial to promote

healthy weight development among 11-13 year olds: The HEalth In Adolescents study. Scand J Public Health. 2010;38(5 Suppl):38-51.

41. James J, Thomas P, Cavan D, Kerr D. Preventing childhood obesity by reducing consumption of carbonated drinks: cluster randomised controlled trial. 2004.

42. James J, Thomas P, Kerr D. Preventing childhood obesity: two year follow-up results from the Christchurch obesity prevention programme in schools (CHOPPS). 2007;335(7623):762.

43. Ma G. Study on comprehensive prevention and control technology of childhood obesity based on dietary nutrition. 2009.

44. Meng L, Xu H, Liu A, van Raaij J, Bemelmans W, Hu X, et al. The costs and cost-effectiveness of a school-based comprehensive intervention study on childhood obesity in China. PLoS One. 2013;8(10):e77971.

45. Rosario R, Araujo A, Oliveira B, Padrao P, Lopes O, Teixeira V, et al. Impact of an intervention through teachers to prevent consumption of low nutrition, energy-dense foods and beverages: A randomized trial. Prev Med. 2013;57(1):20-5.

46. Rosario R, Oliveira B, Araujo A, Lopes O, Padrao P, Moreira A, et al. The impact of an intervention taught by trained teachers on childhood overweight. Int J Environ Res Public Health. 2012;9(4):1355-67.

47. University of Minho. The Impact of an Intervention Taught by Trained Teachers on Childhood BMI z Score. 2011.

53. Research Institute of Child Nutrition Dortmund. Promoting Water Consumption for Prevention of Overweight in School Children in a Controlled Intervention Trial (trinkfit). 2007.

67. Lawlor DA. Active for Life Year 5. 2010.

68. Lawlor DA, Howe LD, Anderson EL, Kipping RR, Campbell R, Wells S, et al. The Active for Life Year 5 (AFLY5) school-based cluster randomised controlled trial: effect on potential mediators. BMC Public Health. 2016;16:68.

69. Lawlor DA, Jago R, Noble SM, Chittleborough CR, Campbell R, Mytton J, et al. The Active for Life Year 5 (AFLY5) school based cluster randomised controlled trial: study protocol for a randomized controlled trial. Trials. 2011;12:181.

70. Lawlor DA, Kipping RR, Anderson EL, Howe LD, Chittleborough CR, Moure-Fernandez A, et al. Active for Life Year 5: a cluster randomised controlled trial of a primary school-based intervention to increase levels of physical activity, decrease sedentary behaviour and improve diet. Public Health Research. 4. Southampton (UK)2016.

71. Lawlor DA, Peters TJ, Howe LD, Noble SM, Kipping RR, Jago R. The Active for Life Year 5(AFLY5) school-based cluster randomised controlled trial protocol: detailed statistical analysis plan. Trials. 2013;14:234.
